# Differentiation of COVID-19 from other emergency infectious disease presentations using whole blood transcriptomics then rapid qPCR: a case-control and observational cohort study

**DOI:** 10.1101/2023.09.03.23294989

**Authors:** Ho Kwong Li, Heather R. Jackson, Luca Miglietta, Dominic Habgood-Coote, Ewurabena Mills, Ravi Mehta, Ali Hamady, Anna Haber, Maisarah Amran, Robert Hammond, Dominique Arancon, Graham S. Cooke, Mahdad Noursadeghi, Peter J.M. Openshaw, Jesus Rodriguez-Manzano, Myrsini Kaforou, Shiranee Sriskandan

## Abstract

**Background:** The overlapping clinical presentations of patients with acute respiratory disease can complicate disease diagnosis. Whilst PCR diagnostic methods to identify SARS-CoV-2 are highly sensitive, they have their shortcomings including false-positive risk and slow turnaround times. Changes in host gene expression can be used to distinguish between disease groups of interest, providing a viable alternative to infectious disease diagnosis.

**Methods:** We interrogated the whole blood gene expression profiles of patients with COVID-19 (n=87), bacterial infections (n=88), viral infections (n=36), and not-infected controls (n=27) to identify a sparse diagnostic signature for distinguishing COVID-19 from other clinically similar infectious and non-infectious conditions. The sparse diagnostic signature underwent validation in a new cohort using reverse transcription quantitative polymerase chain reaction (RT-qPCR) and then underwent further external validation in an independent *in silico* RNA-seq cohort.

**Findings:** We identified a 10-gene signature (*OASL, UBP1, IL1RN, ZNF684, ENTPD7, NFKBIE, CDKN1C, CD44, OTOF, MSR1*) that distinguished COVID-19 from other infectious and non-infectious diseases with an AUC of 87.1% (95% CI: 82.6%-91.7%) in the discovery cohort and 88.7% and 93.6% when evaluated in the RT-qPCR validation, and *in silico* cohorts respectively.

**Interpretation:** Using well-phenotyped samples collected from patients admitted acutely with a spectrum of infectious and non-infectious syndromes, we provide a detailed catalogue of blood gene expression at the time of hospital admission. The findings result in the identification of a 10-gene host diagnostic signature to accurately distinguish COVID-19 from other infection syndromes presenting to hospital. This could be developed into a rapid point-of-care diagnostic test, providing a valuable syndromic diagnostic tool for future early pandemic use.

**Funding:** Imperial COVID fund; NIHR Imperial BRC; UKRI (ISARIC-4C).

**Research in context:** *Evidence before this study:* Rapid diagnosis is fundamental for ensuring that high consequence infections are identified at an early stage, and that correct and timely treatment is started. Pathogen-focused diagnostic tools may not be available early in a pandemic. To determine if host-based syndromic diagnostic tools to identify acute COVID-19 in the emergency setting have been developed, we searched PubMed using the following search terms for all hits between January 2020-July 2023: “COVID19” AND “viral” AND “whole blood” AND (“RNAseq” OR “RNA-Seq” OR “transcriptomic” OR “transcriptome” OR “gene expression”) AND (“signature” OR “diagnosis” OR “classification” OR “classifier”). This returned 16 studies, with two focused on paediatric populations and one focused on an elderly population. A further two studies explored utility of host gene expression in predicting viral infection severity and one study focused on exploring whole blood transcriptome profiles of patients with SARS-CoV-2, however only contrasting them to healthy controls rather than clinically similar disease cohorts. One study demonstrated that metabolomic biomarkers can distinguish COVID-19 and viral infections from other disease groups, and a further study showed that host gene expression (nasopharyngeal swabs and whole blood) differs between patients with COVID-19 and those with influenza, other seasonal coronaviruses, and bacterial sepsis, using classifiers with as few as 20 genes to perform diagnosis. These studies show that acute infection with SARS-CoV-2 can give rise to specific gene expression changes in the host that may differ from those seen in clinically similar infectious or non-infectious presentations. However to date there is no signature that has been adapted to a diagnostic platform, and none has been validated to discriminate SARS-CoV-2 from other infectious syndromes.

*Added value of this study:* Our study provides a unique snapshot of gene expression in a large cohort of well-phenotyped adults at the point of admission to an emergency department with a range of suspected infections including COVID-19. We identified a 10-gene signature, which outperformed common laboratory markers, such as CRP and white cell count for discriminating patients with COVID-19 from those with clinical similar infectious and non-infectious diseases. This signature has been shown to be effective in a completely independent cohort of patients recruited in the United States, as well as in a validation cohort from the emergency department, using a different quantitation platform (RT-qPCR). Taken together, these findings show that acute COVID-19 can be differentiated from other emergency presentations using a sparse combination of host transcripts in blood. The findings allow a gene expression signature to be developed into a rapid point-of-care diagnostic test to differentiate serious COVID-19-like infection from other similar presentations.

*Implications for practice or policy and future research combined with existing evidence.:* PCR-based diagnostic approaches have high sensitivity and specificity to detect SARS-CoV-2 and other viruses in the respiratory tract, however there are many situations where the results may not indicate active disease and can be misleading. Host response-based diagnostics can provide supporting evidence of an active viral infection, and could prove essential in the setting where emerging virus variants elude detection by PCR, or where no PCR diagnostic exists.

## INTRODUCTION

The differential diagnosis of an acutely unwell patient with febrile respiratory syndrome can be challenging, even with the advent of widely available specific respiratory virus PCR testing. As demonstrated by the COVID-19 pandemic, the relevance of a positive viral PCR result can be unclear in settings with high prevalence of SARS-CoV-2, due to prolonged viral RNA shedding, transient oropharyngeal contamination, and high sensitivity of PCR, while viral PCR may be negative in patients with active viral disease due to sample timing, poor sampling, or virus mutation (1–3). A pathogen-specific biomarker of the host response to infection could provide valuable contextual diagnostic information. While clinicians have come to rely upon C-reactive protein (CRP) and procalcitonin to support bacterial diagnoses, there is as yet no accepted biomarker for the host response to viral infection, or COVID-19.

Recently, two host-derived biomarkers of viral infection based on either a sparse host 3-gene expression signature (4) or a single soluble biomarker, ddhC, in serum (5) have been described. Both biomarkers were able to distinguish acute viral infections, including acute COVID-19, from other infectious disease presentations in adults presenting to the emergency department (ED). A number of investigators have described gene expression signatures that differentiate between viral and bacterial infection or non-infectious syndromes (6–9). However, differentiation of acute COVID-19 from all other presentations, including other viral infections, may be more challenging, due to the powerful type 1 interferon response that dominates the host anti-viral immune response in blood. Several groups have reported whole blood gene expression changes in acute COVID-19 compared with other infection syndromes, often combining multiple data sets from separate studies (10–16). To our knowledge, none have validated such a gene expression diagnostic signature in differentiation of acute COVID-19 from other infection presentations.

In this study, we set out to determine if patients with acute COVID-19 can be differentiated from others presenting with suspected infection to the ED, by application of whole blood transcriptomics. Our aim was to undertake a proof of principle study, to determine if a minimal signature to distinguish COVID- 19 from other infections could be derived from a discovery cohort of patients, who had undergone whole blood RNA sequencing (RNA-Seq), and then test this signature in a separate cohort of emergency admissions with suspected infection, using a tractable reverse transcription-quantitative polymerase chain reaction (RT-qPCR) approach, that can be readily adapted to a point of care test. Although our current study is limited to differentiation of COVID-19 from other infectious and inflammatory syndromes, we postulate that identification of future emergent highly pathogenic coronavirus infections might be feasible using such an approach.

## METHODS

### Study design and participants

As part of the BioResource in Adult Infectious Diseases (BioAID), patients who presented with suspected infection to the emergency department (ED) of a major London teaching hospital, were prospectively recruited. At the time of presentation, blood culture and whole blood sampling for RNA extraction was performed, and demographic and clinical data collected as detailed in the BioAID study protocol (17) (UK CRN reference 36653). Ethical approval was granted by the South Central–Oxford C national research ethics committee to obtain deferred consent from participants from whom an RNA specimen had been collected (14/SC/0008 and 19/SC/0116).

### Discovery cohorts

The discovery cohorts comprised BioAID participants recruited September 2014-April 2017 (Discovery A) and those recruited September 2014-June 2020 encompassing the first COVID-19 pandemic wave in the UK (Discovery B); patient selection is described in the Appendix, Supplementary Figure 1. Discovery A patients categorised with bacteremia, non-COVID-19 viral infection, or ‘no infection diagnosed’ were previously reported (4). Discovery B patients were assigned to three categories using the hospital pathology IT system and clinician case note review as follows: bacteremia; non COVID-19 viral infection; and acute COVID-19 (requiring physician confirmation and PCR test). Discovery B included all patients with confirmed viral infections not already included in BioAID A, and the first 100 consented COVID-19 patients recruited into BioAID. Whole-blood RNA sequencing (RNA-seq) of Discovery B was performed for this study. The two cohorts (Discovery A and B) were used to derive a novel gene expression signature to differentiate COVID-19 from other infectious diseases (bacterial and viral) and from patients with suspected sepsis with no infection identified category. Whole blood RNA from 10 consenting normal donors was obtained from a subcollection of the ICHT Tissue Bank (approved by Wales REC3 reference 17/WA/0161).

### Diagnostic signature derivation from RNA-seq

The Discovery dataset was generated by merging the two separate RNA-seq datasets. Discovery datasets A and B are available on ArrayExpress (E-MTAB-10527 and E-MTAB-13307 respectively) Processing and analysis of RNA-seq data are described in the Appendix. In brief, whole blood transcriptome profiles from individuals with COVID-19 were contrasted to those from individuals with bacterial infections, viral infections, and no-infection unwell controls to identify a sparse combination of host genes that could distinguish COVID-19 from these disease groups. Gene expression signatures were identified using Forward Selection - Partial Least Squares (FS-PLS) (18).

### Validation of the diagnostic gene signature using RT-qPCR in patients presenting to the ED

The diagnostic gene signature derived was then validated in RNA samples from a separate, combined validation cohort, using RT-qPCR to quantify expression of the genes in the signature, with glyceraldehyde 3-phosphate dehydrogenase (*GAPDH*) as a housekeeping gene. The cohort comprised 228 BioAID patients prospectively recruited over 6 months with suspected infection pre-COVID-19, as previously described (4), plus 50 BioAID patients with PCR-confirmed acute COVID-19, summarised in the Appendix (Supplementary figure S2). All patients were unvaccinated against COVID-19. RT-qPCR workflow required reverse transcription, pre-amplification, on-chip gene expression, and data analysis (supplementary appendix). The performance of the host gene signature to classify COVID-19 against all other groups was tested. The cycle threshold (CT) values of genes measured were combined into disease risk scores (DRS) for each patient, using the simple DRS which was calculated through subtracting the total expression of genes expected to be downregulated in COVID-19 from the total expression of genes expected to be upregulated in COVID-19, using the directions determined from the model weights estimated in the discovery dataset, as follows:

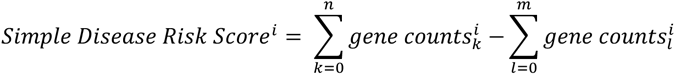

Where *i* represents each sample; *n* represents each gene included in the signature that increases in COVID-19 and *m* represents each gene included in the signature that decreases in COVID-19.

A retrained DRS was also used, which was calculated through multiplying each gene’s CT values by logistic regression model coefficients retrained on the RT-qPCR dataset (retrained model weights) contrasting COVID-19 vs. bacterial infections, viral infections, and not infected (the groups included in the discovery stage). The retrained DRS represents the ceiling of performance as the weights are refitted to each dataset.

The performance of the signature was contrasted to performance of clinical variables typically used clinically, C-reactive protein (CRP) and leukocyte counts (WCC).

### External validation of the diagnostic gene signature in silico

A publicly available RNA-seq dataset (McClain et al, 2021) (19) that included patients with COVID- 19; bacterial infections, influenza, seasonal coronaviruses, and healthy controls was used for *in silico* validation of the diagnostic gene signature (Supplementary Appendix). AUCs were calculated using the simple DRS using the expected direction of change from the discovery stage, as well as using the retrained DRS with model weights retrained through logistic regression models contrasting COVID- 19 to all other groups (bacterial, influenza and other coronaviruses). The simple DRS was calculated using the equation described above.

## Results

### Clinical characteristics of combined discovery cohorts

Discovery cohort A, which has previously been described, comprised RNA from patients with confirmed bacteraemia (n=60), non-COVID-19 viral infection (n=30), no final microbiological diagnosis (inferred to not be infected, n=30) (4). Discovery B comprised patients with acute COVID- 19 patients (n=100), non-COVID-19 viral infection (n=9), bacteraemia (n=31), and healthy controls (n=10) (Supplementary Figure 1).

Patients in the bacterial and COVID-19 groups were older than other groups, although a greater number of bacterial patients had comorbidities in comparison to COVID-19 patients. CRP was significantly lower in the not-infected group compared with either the bacterial or COVID-19 groups **(Table 1),** however notably there was no difference when comparing COVID-19 and bacterial groups with one another.

**Table 1.** Clinical characteristics of the combined Discovery cohorts A and B.

|  | Discovery A & B RNA-Seq Cohort |  |  |  |
| --- | --- | --- | --- | --- |
| Characteristic | Bacterial<br>(n = 88) | Viral<br>(n = 36) | Not-infected<sup>#</sup><br>(n = 27) | COVID-19<br>(n=87) |
| Age - years<sup>α</sup> |  |  |  |  |
| Median (IQR) | 69.0 (57.3-79.8) | 44.0 (27.8-56.5) | 48.0 (31.0-70.0) | 66.0 (53.0-79.0) |
| Range | 25.0-95.0 | 19.0-89.0 | 19.0-96.0 | 23.0-97.0 |
| Sex - frequency (%) |  |  |  |  |
| Male sex | 44 (50.0) | 21 (58.3) | 17 (63.0) | 50 (57.5) |
| Comorbidities<sup>β</sup> (%) |  |  |  |  |
| No | 35 (39.8) | 25 (69.4) | 11 (40.7) | 64 (73.6) |
| Yes | 53 (60.2) | 11 (30.6) | 16 (59.3) | 23 (26.4) |
| Admission temperature -<br>Celsius<sup>γ*</sup> |  |  |  |  |
| Median (IQR) | 38.1 (37.0-38.9) | 38.4 (37.8-38.9) | 37.0 (36.5-37.3) | 37.9 (36.9-38.2) |
| Range | 35.5-40.8 | 35.4-39.8 | 35.6-37.9 | 33.4-39.7 |
| Admission blood tests -<br>median (IQR) |  |  |  |  |
| Leukocytes<sup>δ</sup> (10<sup>9</sup>/L) | 12.4 (8.3-15.9) | 7.3 (5.1-9.3) | 8.9 (6.3-12.1) | 8.0 (6.2-9.6) |
| Neutrophils<sup>δ</sup> (10<sup>9</sup>/L) | 10.7 (6.6-14.2) | 5.4 (3.9-7.1) | 6.1 (4.5-8.3) | 6.2 (4.3-8.2) |
| Lymphocytes<sup>ε</sup> (10<sup>9</sup>/L) | 0.8 (0.5-1.2) | 0.8 (0.6-1.1) | 1.6 (0.9-2.5) | 0.8 (0.6-1.3) |
| CRP<sup>φ*</sup> (mg/L) | 105.0 (49.3-204.0) | 33.5 (16.6-75.7) | 3.4 (1.3-14.3) | 104.0 (59.8-195.0) |
| Inpatient Mortality - (%) |  |  |  |  |
| Died | 24 (27.3) | 2 (5.6) | 4 (14.8) | 25 (28.7) |

<sup>#</sup>Two cases in the Not-infected group were separate admissions from one patient.
<sup>α</sup>Significant difference between Bacterial vs. Viral; Bacterial vs. Not-infected; Viral vs. COVID-19; and Not-infected vs. COVID-19 groups where $p < 0.001$ (Tukey's multiple comparison test).
<sup>β</sup>Significant difference between Bacterial vs. COVID-19 groups.
<sup>γ</sup>Significant difference between Bacterial vs. Not-infected, and Viral vs. Not-infected groups.
<sup>δ</sup>Significant difference between Bacterial vs. Viral, and Bacterial vs. COVID-19 groups.
<sup>ε</sup>Significant difference between Bacterial vs. Not-infected groups.
<sup>φ</sup>Significant difference between Bacterial vs. Not-infected; Viral vs. COVID-19, and Not-infected vs. COVID-19 groups.
Kruskal-Wallis (Dunn's multiple comparison test) was used for all comparisons unless otherwise stated, significance is where $p < 0.001$
\*Tests missing/not done at time of admission: Seventeen temperature readings (Bacterial group, 9; Viral group, 2; Not-infected group 6); twenty-two CRP results (Bacterial group, 15; Viral group, 3; Not-infected group, 3; COVID-19, 1)

### Signature discovery to identify COVID-19 from all other patients

There were 18,247 quantified genes in the normalised, merged dataset, initially appraised by principal component analysis. (Supplementary Figure S3). We first compared COVID-19 patients with each of the other disease groups and applied FS-PLS (18) to derive small 2-gene and 4-gene signatures to distinguish the groups (Supplementary Table S1 and Supplementary Figure S4). This yielded two 2- gene signatures (COVID-19 vs bacterial; COVID-19 vs viral) and two 4-gene signatures (COVID-19 vs bacterial + viral + non-infected; COVID-19 vs bacterial + viral). The ten unique genes that comprised these small signatures were then combined into a single model to explore their combined performance (Supplementary Figure S5) **(Table 2**). We evaluated the ability of the ten gene-signature to distinguish COVID-19 patients from all the non-COVID-19 patients combined, by calculating the DRS with weights obtained from a logistic regression model. The 10-gene signature achieved an AUC of 87.1% (95% CI: 82.6%-91.7%) when distinguishing COVID-19 from all non-COVID-19 disease groups, when applied to the original discovery dataset **(Figure 1).**

**Figure 1.**
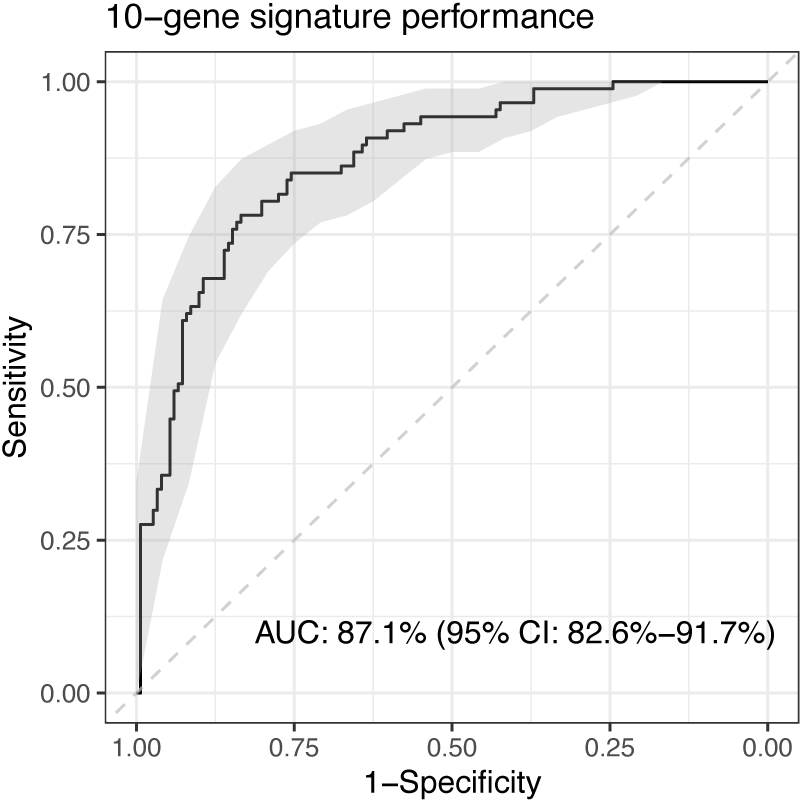
Performance of the pooled 10-gene signature in the discovery RNA-seq dataset. The 10 genes were identified from FS-PLS models contrasting COVID-19 to bacterial infections, viral infections and non-infected patients individually and in combination. Performance of 2-gene and 4- gene signatures that contribute to the 10-gene signature is in Supplementary Appendix.

**Table 2.**
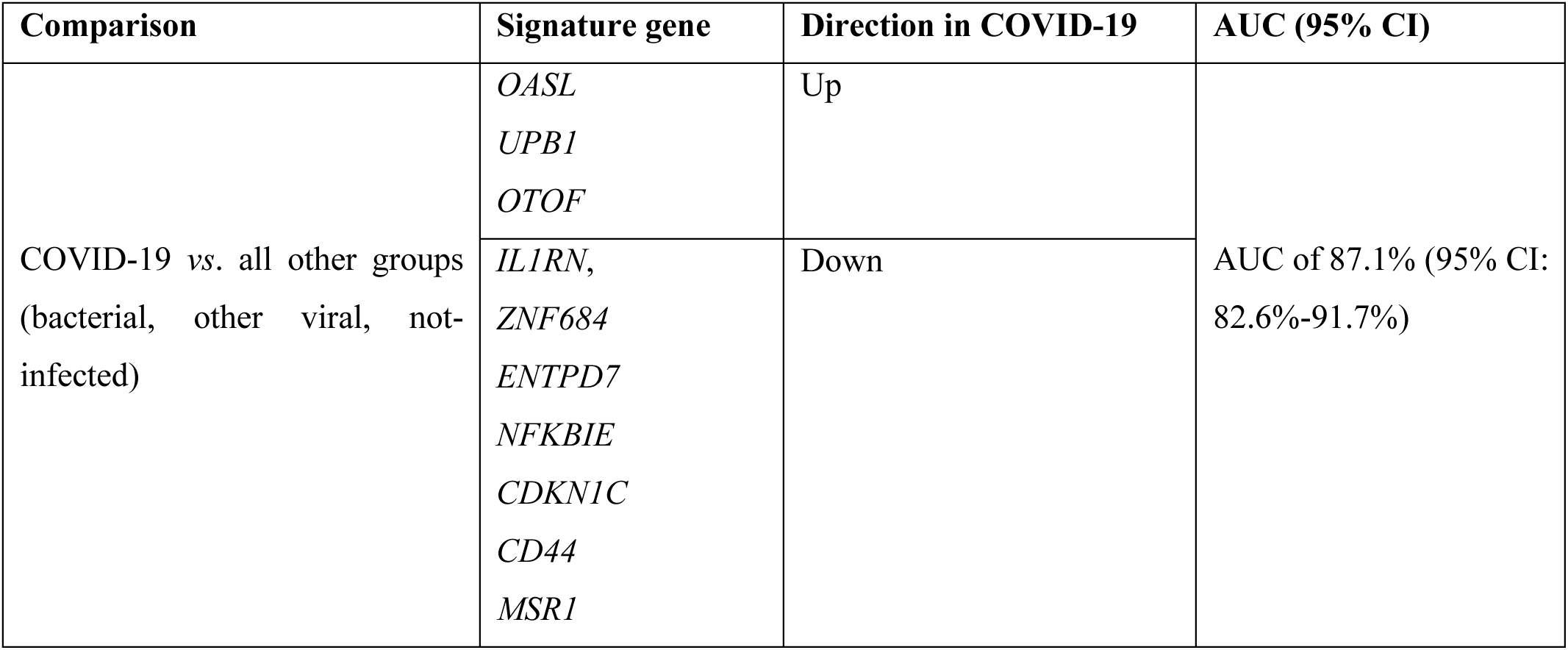
Components of ten gene signature to distinguish acute COVID-19 from all other infectious presentations.

### Validation of the 10-gene signature by RT-qPCR in distinct RNA samples

The ability of the 10-gene signature to identify COVID-19 was then explored using a combined validation cohort **(Table 3)** comprising 167 RNA samples from a ‘pre-COVID-19’ validation cohort (consecutive ‘real life’ suspected infection admissions with a range of final diagnoses) and 37 RNA samples from a COVID-19 validation cohort that were suitable for RT-qPCR analysis (Supplementary Figure S2). RT-qPCR revealed *CD44*, *MSR1*, *ENTPD7*, *NFKBIE*, *CDKN1C* and *IL1RN* levels to be significantly lower in COVID-19 patients compared to those with bacterial infection and not infected (Supplementary Figure S6). *NFKBIE*, *CDKN1C* and *IL1RN* levels were significantly lower in COVID- 19 compared to viral infection. *OTOF* levels were significantly higher in COVID-19 compared to bacterial infection and not infected.

**Table 3.** Clinical characteristics of the combined pre-COVID-19 and COVID-19 RNA validation cohorts tested using RT-qPCR.

|  | COVID-19 CASE CONTROL VALIDATION COHORT |  |  |  |  |  |
| --- | --- | --- | --- | --- | --- | --- |
| CHARACTERISTIC | Bacterial (n = 28) | Viral (n = 4) | Probable / Indeterminate (n = 112) | Not-infected (n = 20) | Other Infection (n = 3) | COVID-19 (n = 37) |
| Age - years |  |  |  |  |  |  |
| Median (IQR) | 66.5 (55.3-79.0) | 56.0 (30.8-73.0) | 62.5 (42.3-73.5) | 58.5 (45.8-69.8) | 74.0 (-) | 66.0 (49.5-77.0) |
| Range | 20.0-91.0 | 23.0-78.0 | 18.0-91.0 | 21.0-94.0 | 70.0-84.0 | 24.0-88.0 |
| Gender - frequency (%) |  |  |  |  |  |  |
| Male sex | 10 (35.7) | 3 (75.0) | 64 (57.1) | 9 (45.0) | 2 (66.7) | 22 (59.5) |
| Comorbidities (%) |  |  |  |  |  |  |
| No | 7 (25.0) | 2 (50.0) | 39 (34.8) | 11 (55.0) | 2 (66.7) | 16 (43.2) |
| Yes | 21 (75.0) | 2 (50.0) | 73 (65.2) | 9 (45.0) | 1 (33.3) | 21 (56.8) |
| Admission temperature - Celsius<sup>κ*</sup> |  |  |  |  |  |  |
| Median (IQR) | 38.0 (36.9-39.0) | 37.4 (36.6-38.2) | 37.4 (36.6-38.1) | 36.4 (35.6-37.0) | 38.1 (-) | 37.5 (36.8-38.2) |
| Range | 32.0-39.4 | 36.5-38.2 | 31.3-39.9 | 33.3-38.0 | 37.9-38.2 | 35.7-40.4 |
| Admission blood tests - median (IQR) |  |  |  |  |  |  |
| Leukocytes<sup>λ*</sup> (10<sup>9</sup>/L) | 13.8 (9.7-19.2) | 8.2 (3.8-8.7) | 10.7 (8.1-14.7) | 9.3 (6.8-13.7) | 11.4 (-) | 6.9 (5.0-8.8) |
| Neutrophils<sup>λ*</sup> (10<sup>9</sup>/L) | 10.8 (7.6-17.0) | 6.5 (3.0-6.8) | 8.4 (5.4-12.1) | 7.2 (4.1-10.6) | 10.5 (-) | 5.4 (3.8-7.5) |
| Lymphocytes<sup>*</sup> (10<sup>9</sup>/L) | 0.8 (0.4-1.4) | 0.8 (0.3-1.4) | 1.1 (0.7-1.6) | 1.9 (0.8-2.5) | 0.6 (-) | 0.8 (0.6-1.1) |
| CRP<sup>μ</sup> (mg/L) | 176.0 (89.6-265.0) | 55.0 (29.0-113.0) | 63.5 (15.7-184.0) | 6.4 (4.7-51.2) | 131.0 (-) | 103.0 (60.5-139.0) |
| Inpatient mortality - frequency (%) |  |  |  |  |  |  |
| Died | 8 (28.6) | 0 (0.0) | 3 (2.7) | 2 (10.0) | 0 (0.0) | 9 (24.3) |

<sup>κ</sup> Significant difference Bacterial vs. Not-infected groups
<sup>λ</sup> Significant difference Bacterial vs. COVID-19, and Indeterminate vs. COVID-19 groups
<sup>μ</sup> Significant difference between Bacterial vs. Not-infected, and Not-infected vs. COVID-19 groups.
Kruskal-Wallis (Dunn's multiple comparison test) was used for all comparisons, significance is where $p < 0.001$
\*Tests missing/not done at time of admission: Four temperature readings (Not-infected group, 2; Other Infection group, 1; COVID-19 group 1); one WCC/neutrophil/lymphocyte result (COVID-19 group); sixteen CRP results (Bacterial group, 1; Indeterminate group, 9; Not-infected group, 4; Other Infection group, 1; COVID-19, 1)

When COVID-19 was contrasted against all comparator groups combined, the 10-gene signature achieved an AUC of 83.2% (95% CI: 76.3%-90%) using the simple DRS, which improved to 88.7% (95% CI: 84.2%-93.1%) when the model weights were retrained (**Figure 2A**). The ability of the 10- gene signature to distinguish COVID-19 from all other ‘definite infections’, a group composed of bacterial, viral, and other infections, was explored, yielding an AUC of 90.5% (95% CI: 83.3%-97.7%) with the simple DRS, and 96.9% (95% CI: 93.7%-100%) with retrained weights (**Figure 2B**). When COVID-19 was contrasted against ‘probable/indeterminate infections’, the 10-gene signature achieved an AUC of 79.8% (95% CI: 71.9%-87.6%) which improved to 84.7% (95% CI: 78.7%-90.7%) with retrained weights (**Figure 2C).** Performance against both of these groups combined is shown in the Appendix (Supplementary Figure S7). Finally, the performance of the signature in distinguishing COVID-19 from the ‘not-infected’ phenotypic group led to a performance of 89.2% (95% CI: 79.6%- 98.8%) using the simple DRS, and 96.1% (95% CI: 91.3%-100%) with retrained model weights (**Figure 2D)**.

**Figure 2:**
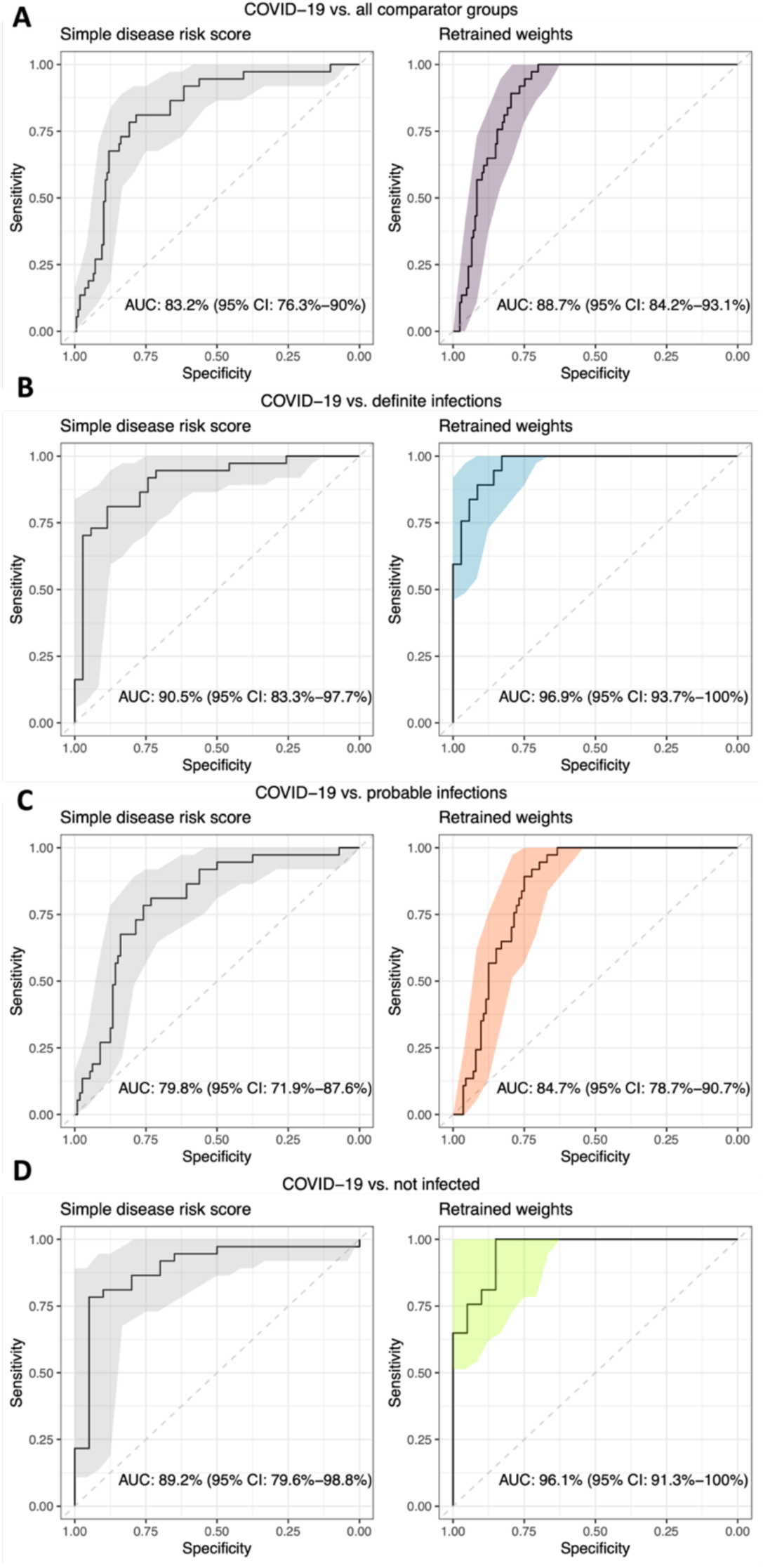
ROC curves showing the performance of the 10-gene signature in the RT-qPCR validation dataset in distinguishing between A) COVID-19 vs. all groups, B) COVID-19 vs. definite infections (bacterial, viral, mixed), C) COVID-19 vs. probable infections (probable bacterial, probable viral, indeterminate), D) COVID-19 vs. not infected. ROC curves and AUCs shown in plots on the left-hand side panel are calculated using the simple disease risk score (DRS), whilst those on the right-hand side panel are calculated using retrained model weights.

The performance of the 10-gene signature was contrasted to the performance of clinical variables typically used: C-reactive protein (CRP) and leukocyte cell counts (WCC) (**Figure 3).** Importantly, when comparing COVID-19 with all comparator groups, CRP had an AUC of 57.5% (95% CI: 49.3%- 65.7%) and WCC had an AUC of 77.9% (95% CI: 71.2%-84.6%) in contrast to an AUC of 83.2-88.7% when using the 10-gene signature (**Figure 3A**). For COVID-19 *vs*. definite infections, and COVID-19 *vs*. probable infections, the 10-gene signature outperformed CRP and leukocyte count **(Figure 3B & C).** The 10-gene signature outperformed CRP and WCC for all comparisons except where retrained weights were used in the COVID-19 *vs* not-infected group (**Figure 3D**), for which CRP outperformed the 10-gene signature.

**Figure 3.**
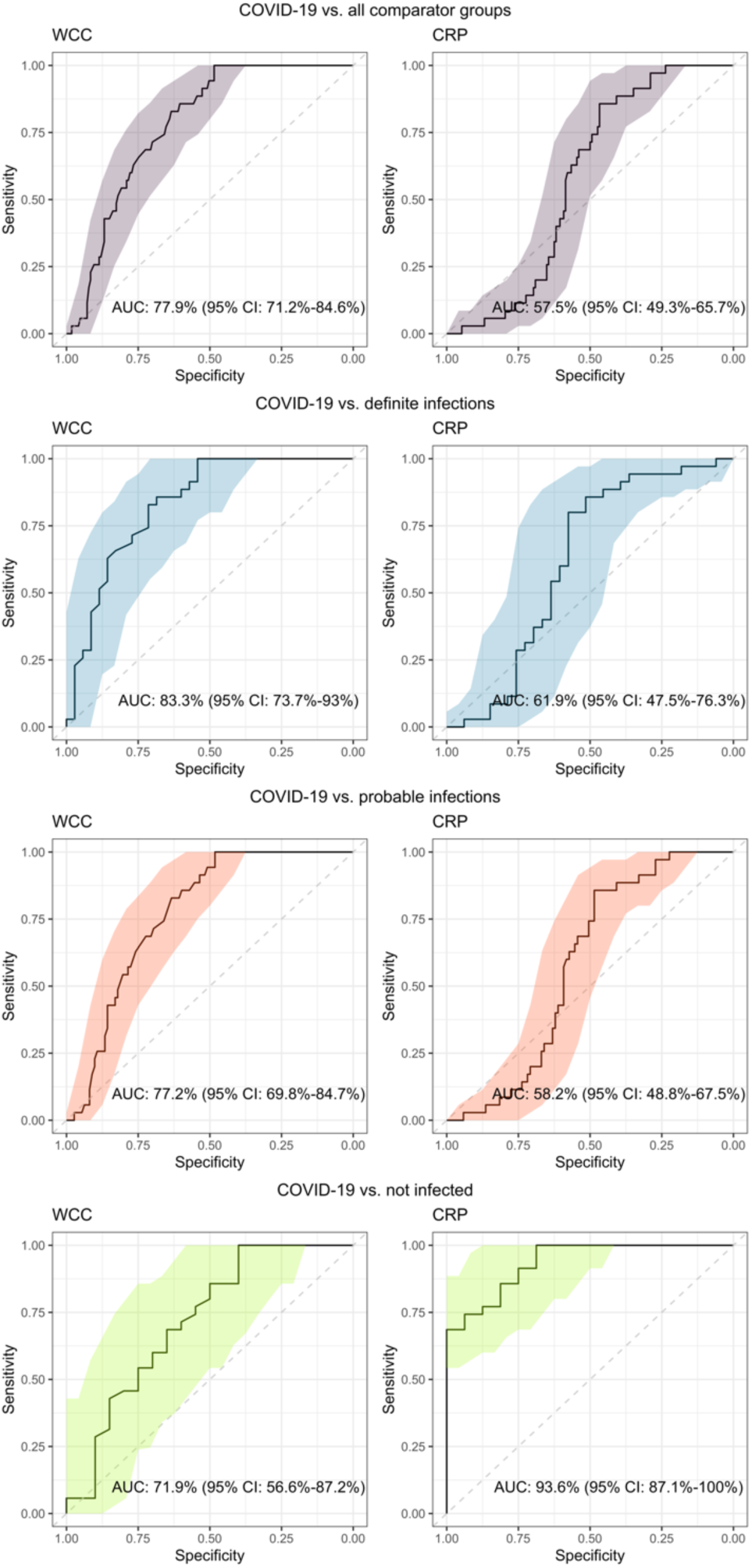
Performance of routine laboratory biomarkers CRP and leukocyte count (WCC) when comparing (A) COVID-19 and all comparator groups in the validation cohorts; (B) COVID-19 and the definite infection groups; (C) COVID-19 and probable infection groups and (D), COVID-19 and the not infected group.

### External validation of 10-gene signature in silico using published dataset

An independent, publicly available RNA-seq validation dataset from McClain et al. (19) was downloaded from Gene Expression Omnibus; samples from COVID-19 patients with early (≤10 days, n=19) and middle symptom duration (11–21 days, n=36), bacterial infections (n=23), viral infections (influenza n=17 and other seasonal coronaviruses n=57) and healthy controls (n=22) were used for validation of the 10-gene signature.

When ‘early’ COVID-19 samples were contrasted against all other infections combined, the 10-gene signature achieved an AUC of 82.4% (95% CI: 73.4%-91.3%) which increased to 93.6% (95% CI: 86.2%-100%) when the model weights were retrained using a generalised logistic regression model contrasting COVID-19 to bacterial and viral infections **(Figure 4A).** The performance of the 10-gene signature was further evaluated through contrasting early COVID-19 to bacterial and viral infections separately. The 10-gene signature achieved an AUC of 99.5% (95% CI: 98.5%-100%) in distinguishing COVID-19 from bacterial infections using the simple DRS, which decreased to 93.6% (95% CI: 86.6%-100%; **Figure 4B)** following retraining of model weights. The AUC for the 10-gene signature in distinguishing between COVID-19 and other viral infections was 77.1% (95% CI: 65.9%- 88.4%) with the simple DRS, increasing to 93.6% (95% CI: 85.5%-100%) following retraining of model weights (**Figure 4C).** The performance for the 10-gene signature was lower when middle COVID-19 samples were included in the AUROC calculations, for calculations with both the simple DRS and retrained weights (Supplementary Figure S8).

**Figure 4.**
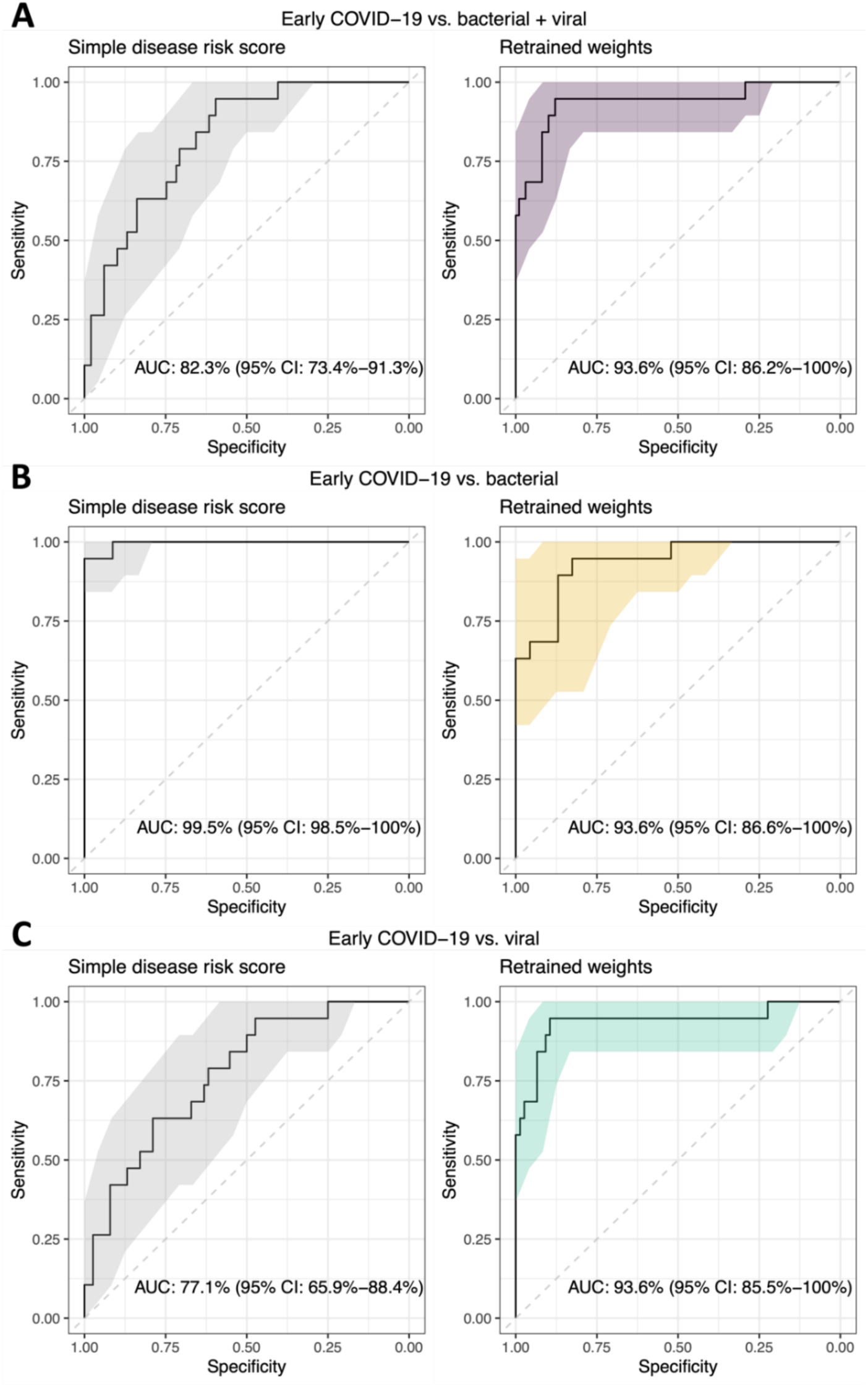
ROC curves showing the performance of the 10-gene signature in the *in-silico* RNA-seq validation dataset (19) in distinguishing between A) COVID-19 vs. all groups, B) COVID-19 vs. bacterial infections, C) COVID-19 vs. viral infections. ROC curves and AUCs shown in plots on the left-hand side panel are calculated using the simple DRS, whilst those on the right-hand side panel are calculated using retrained model weights.

### Gene expression in patients with and without infection

As adult emergency presentations may frequently masquerade as infection, our final analysis identified genes differentially expressed between patients who were originally recruited into BioAID (due to suspicion of infection) but who had no evidence of infection at discharge, and those in any of the definite infection categories (bacterial, viral, and COVID-19). *SLC24A2* and *CD1E* were previously identified as over-expressed in patients without infection vs. patients with infection using Discovery A (4). Using an adjusted *p*-value threshold of < 0.0001, and an absolute log2 fold-change (LFC) threshold of > 1.5, an additional 29 upregulated genes were identified in infected samples vs. not-infected patients in the merged dataset that included COVID-19 patients (Supplementary Table S2).

## DISCUSSION

Li et al previously identified a minimal 3-gene signature that could distinguish adults with acute viral infections (including COVID-19) from others presenting with acute infections, and showed that categorisation could be achieved with a simple RT-qPCR-based diagnostic test (4). In this report we show that it is possible to use an expanded host gene expression signature to distinguish patients with acute COVID-19 from all other patients presenting to the emergency department with infection syndromes. Coupled with other routinely applied diagnostic tests, such a signature could provide rapid and valuable contextual information to clinicians, much in the same way that CRP is used already. The 10-gene signature displayed good performance in differentiating COVID-19 from other infectious diseases, in addition to differentiation of ‘not infected’ patients masquerading as infection, a common scenario in adult medicine. The signature has translated across platforms (from RNA-seq to RT-qPCR) and also across cohorts: discovery and initial RT-qPCR validation used samples recruited from the UK BioAID study (4, 17), while the *in silico* validation used independent data from the United States (19). When used to distinguish COVID-19 from the large and pleiotropic group of all other febrile presentations to the ED, the 10-gene signature yielded AUCs of up to 87.1%, 88.7%, and 93.6% when evaluated in the discovery, RT-qPCR validation, and *in silico* cohorts respectively.

Host blood transcriptomic signatures are well-recognised for their ability to differentiate infections based on aetiology, severity and even symptomatology (6–16, 20–25). Although there are useful existing gene expression signatures that can differential acute viral infections from bacterial sepsis, more granular differentiation of COVID-19 from other viral infections such as influenza, RSV, or rhinovirus may be crucial to ongoing patient management, both in terms of infection control decisions as well as treatment. Furthermore, a reliable host gene signature that differentiates COVID-19 from other viral or bacterial infections as well as cases of not-infectious syndromes masquerading as infection, would for example, greatly support acute clinicians in prescribing anti-inflammatory therapy indicated in one infection but contraindicated in the other.

Although many other investigators have compared whole blood transcriptomic data obtained from COVID-19 with data obtained from patients with other infections (10–16), we are not aware of any that have derived a gene expression signature that has been validated in a fresh cohort of patients using different methodology. A major strength of our work is that all samples were obtained at the point of admission to hospital, at the time of the very first blood tests being conducted, which is precisely the time that a diagnostic would be of greatest use. This also meant that patients had not received any treatments that might complicate any gene expression patterns (20). Our ability to compare COVID- 19 with the wide range of heterogeneous presentations to the emergency department is unique: BioAID has been continuously recruiting RNA samples from patients presenting to the emergency department with suspected infection since 2014, and continued to recruit during the first and subsequent waves of COVID-19. The samples from patients with subsequently confirmed bacterial, viral, or no infection are therefore directly comparable and do not suffer from the technical challenges often faced when comparing cohorts recruited from different studies. Finally, we have shown that the signatures that can be derived from gene expression studies can be translated into tractable and potentially economic bedside molecular blood tests.(21)

A number of investigators have applied whole blood transcriptomics to identify predictive scores for deterioration in COVID-19 (20, 22–26). We did not attempt this in our cohort as the number of COVID- 19 patient RNA samples that successfully underwent RNA-seq was relatively small (n=88) and all stemmed from the first (Wuhan) wave of COVID-19, prior to specific treatments being available, with limited access to intensive care or non-invasive ventilation, and at a time when inpatient mortality was very high, just under 30%. To investigate and then validate such a score would have been challenging since treatments for COVID-19 developed rapidly after recruitment of our discovery cohort, although notably mortality from COVID-19 was still 24.3% in the validation cohort. Interestingly although COVID-19 mortality in our discovery cohort was similar to the bacteremia group (28.7% and 27.3% respectively) it was notable that comorbidities were far more frequent among bacteremia patients (60.2%) than the COVID-19 patients (26.4%), a difference that was maintained, albeit to a lesser degree, in the validation cohorts, underlining the devastating impact of the first wave of COVID-19 on otherwise healthy UK patients.

The COVID-19 pandemic and other recent infectious disease events (27) provide a stark reminder that host-based biomarkers could aid diagnosis when faced with emerging infectious syndromes such as rapidly progressive pneumonia. The possibility of SARS reappearance, together with the emergence of avian influenza (H5N1) and pandemic influenza (H1N1) have no doubt motivated improvements in respiratory viral molecular diagnostic testing, which have been enthusiastically adopted in emergency departments. Nonetheless, the value of complementary blood tests to support positive and negative diagnoses should not be under-estimated. Rapid diagnostics for bacterial disease have lagged behind viral infections, meaning that diagnosis in the ED can be influenced or skewed by an uneven use of rapid tests. The potential for false reassurance was highlighted during the winter 2022 resurgence of invasive group A streptococcal infection, where over one quarter of children subsequently found to have invasive group A streptococcal infection, a disease of high mortality, had PCR evidence of a respiratory virus infection (28). A host-based syndromic test to highlight ‘predominant’ viral or bacterial infection in such cases would be greatly valued. Whether a host-based gene expression signature could distinguish active viral infection from convalescent infection will require further work; potentially exploiting human challenge models or longitudinal studies in other cohorts. Certainly it appears that presymptomatic viral infection might also be detectable via gene expression (29).

Our study was limited by the number of new samples that could practically be sequenced in the time frame available in the early stages of the COVID-19 pandemic (n=150). Nonetheless, the first 100 samples from consented SARS CoV-2-positive patients were subject to RNA-seq, providing a valuable publicly-available resource for future scientific research, while a further 50 samples were used to evaluate the 10-gene signature using RT-qPCR. Although our primary aim was to identify a host gene expression signature that could differentiate COVID-19 from other infections, we acknowledge that some of our findings could be confounded by gene expression changes relating to disease severity. Finally, the phenotype of COVID-19 has changed considerably since inception of this study, and it is likely that the pattern of gene expression will also have changed in a vaccinated adult population, and potentially in response to newer variants of SARS-CoV-2.

Host-derived biomarkers of viral and bacterial infection could be of great value in the early stages of a pandemic (5, 30). Our results serve as proof of principle, that the described 10-gene signature can differentiate a severe coronavirus infection such as COVID-19 from other infectious presentations and presentations masquerading as infection in adults. We posit that such a test would be a valuable asset in the event of a future coronavirus threat and recommend that such tests are developed in advance of the next coronavirus pandemic.

## Supporting information

Supplementary Methods and Results

## Acknowledgements

The authors would like to acknowledge the NIHR Biomedical Research Centre (BRC) at Imperial College London, the NIHR BRC Leonard and Dora Colebrook AMR Laboratory, the Imperial NIHR Health Protection Unit in Healthcare-associated infection and AMR for their support of BioAID as well as the NIHR Clinical Research Network (CRN), the Imperial College Healthcare NHS Trust Research nursing team and innumerable volunteers who contributed to patient recruitment and data collection. HKL and RM are UKRI (MRC) Research Training Fellows. MN acknowledges support from the Wellcome Trust (207511/Z/17/Z) and the NIHR Biomedical Research Centre at UCL and UCLH. HRJ acknowledges support from the Wellcome Trust (4-year PhD programme, grant number 215214/Z/19/Z). JRM and MK acknowledge the Rosetrees Trust (PGL22/100101). JRM is affiliated with the Department of Health and Social Care (DHSC) Centre for Antimicrobial Optimisation at Imperial College, London.

## Author contributions

Conceptualisation: SS, HKL, MK; Study design: MK, JRM, SS; Data collection: HKL, LM, EM, RM, AH, MA, RH, DA; Formal analysis: HRJ, MK, DH-C, HKL, AH; Methodology: MN, MK, JRM; Data visualisation: HJ, HKL; Data curation: HKL, RM, HRJ, DH-C, LM, AH; Funding acquisition and supervision: SS, GC, MK, JRM, PO; Writing original draft: HKL, HJ, SS; Review and editing: all authors; Non identifiable data verification: HJ, DC, LM; Identifiable data verification: HKL, RM. All authors had access to all of the non-identifiable data used in the study and support the decision to submit for publication.

## Data Availability Statement

De-identified participant level data and metadata leading to signature discovery are available on ArrayExpress (E-MTAB-10527 and E-MTAB-13307 respectively). BioAID study protocol and other materials are available at https://imperialbrc.nihr.ac.uk/2020/01/28/bioaid/. De-identified participant level data from BioAID patients can be made available to investigators whose proposed use of the data has been approved by the BioAID governance review committee.

