## Supplementary Methods and Results for "Differentiation of COVID-19 from other emergency infectious disease presentations using whole blood transcriptomics then rapid qPCR: a case-control and observational cohort study"


| Contents of Appendix | Page numbers |
|---|---|
| **Supplementary Methods** | **2-5** |
| Figure S1 | **2** |
| Figure S2 | **4** |
| **Supplementary Results** | **6-18** |
| Figure S3 | **6** |
| Table S1 | **7** |
| Figure S4 | **8** |
| Figure S5 | **9** |
| Figure S6 | **10-11** |
| Figure S7 | **12** |
| Figure S8 | **13** |
| Table S2 | **14-18** |
| **References for Appendix** | **19-20** |

### Supplementary Methods

#### Discovery cohorts for RNA sequencing analysis

Discovery A has previously been described (1) and comprises participants with either a confirmed bacterial infection (bacteraemia), a confirmed viral infection, or a 'not-infected' syndrome with an alternative final diagnosis. As the purpose of the current study was to distinguish COVID-19 from all types of infection presentation, RNA-seq from 3 patients with mixed infections (non-COVID-19) that had been previously excluded from Discovery A (4) were included in the current study. Discovery B were participants who were diagnosed with either a confirmed bacteraemia, or a confirmed viral infection, like Discovery A, but also included a SARS-CoV-2 PCR-positive acute COVID-19 group, and a healthy control group.

**Supplementary Figure S1**: **Study profile of Discovery cohorts A and B.**

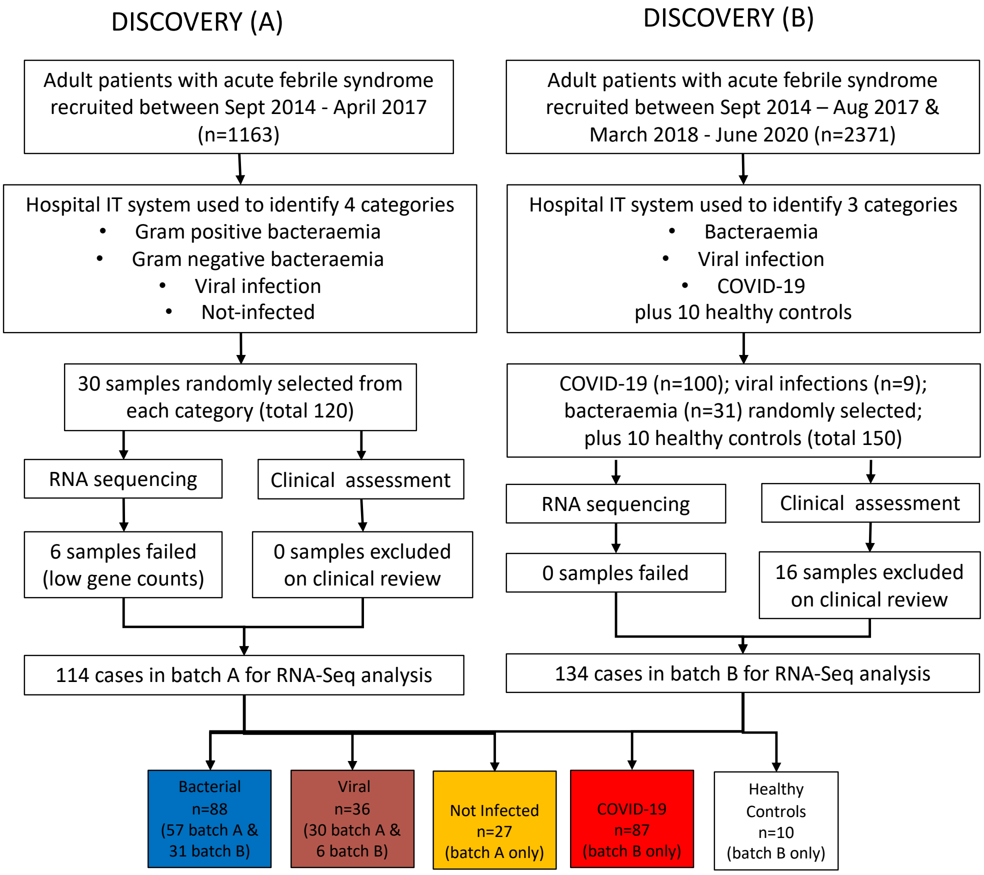


**Supplementary Figure S1**: Whole blood RNA from Discovery cohort B underwent *de novo* RNA sequencing (RNA-seq) for this study; samples were excluded on clinical review when date of pathology result from IT system did not coincide with the BioAID admission date in clinical record. All COVID-19 patients required a clinical presentation consistent with COVID-19 and a positive SARS-CoV2 PCR at the time of admission. Discovery B was drawn from the entire BioAID cohort, excluding a single 6 month period when all participants were used for validation studies.

**Whole blood transcriptomic sequencing for Discovery B**

Transcriptomic sequencing procedures for Discovery B were the same as those for Discovery A. Whole blood was collected at the time of recruitment in Tempus Blood RNA tubes and total RNA was isolated with the Tempus$^{TM}$ Spin RNA Isolation Kit (ThermoFisher Scientific) according to the manufacturer's instructions. RNA samples were stored at −80 °C until further analysis. After additional DNAse treatment, library preparation and sequencing of 30 million 100bp, paired end reads were conducted using the Illumina's TruSeq® RNA Sample Preparation Kit; ribosomal and globin RNA depletion was performed using the Illumina Ribo-Zero Gold kit and HiSeq 4000 at The Wellcome Centre for Human Genetics in Oxford UK. RNA samples for RT-qPCR were quantified using Qubit™ 4 Fluorometer (ThermoFisher Scientific) and NanoDrop® ND-1000 spectrophotometer (ThermoFisher Scientific) to ensure the integrity and quality of the RNA (260:280 ratio between 1.5 and 1.8). Fastq files were trimmed using trimmomatic (2). Quality control was performed using FastQC (3) and MultiQC (4), mapping to the human genome was carried out using STAR (5), and featureCounts (6) was used for read counting. Ensembl Version 89 GCh38 genome and annotation were used (7).

**Diagnostic signature derivation from RNA-seq**

Pre-processing steps were applied to the two datasets (Discovery A and B) separately prior to merging them. Six samples in Discovery A (1) were excluded due to technical factors (failing quality control, low RNA sequencing count). Samples in Discovery A were sequenced on two plates. DESeq2 (8) was applied to the Discovery A counts from bacterial and viral samples to identify significantly differentially expressed (SDE) genes between the two plates. Sex and disease group (bacterial or viral) were also included in the model. Genes with Benjamini-Hochberg (BH) (9) adjusted $p$-values < 0.001 were removed. Genes missing in all samples in either dataset in addition to ribosomal genes were removed from both datasets. Following filtering of genes, Discovery A and B were merged, using only genes present in both datasets. ComBat-seq (10) used to remove batch effects between Discovery A and B, in addition to the plate effects from the two plates in Discovery A. Normalisation was performed using DESeq2 (8) using default parameters. Normalised genes with fewer than three samples with a normalised read count greater than 10 were removed. Principal component analysis (PCA) was performed on the normalised, merged counts.

Differential expression analysis was performed using DESeq2 (8) on the following comparisons: COVID-19 *vs*. bacterial; COVID-19 *vs*. viral; COVID-19 *vs*. not-infected; COVID-19 *vs*. bacterial and viral combined; and COVID-19 *vs*. bacterial, viral, and not-infected combined. Genes were considered significantly differentially expressed (SDE) if they had BH adjusted $p$-values < 0.05. The $\log_2$-transformed normalised, merged counts of the SDE genes were put forward for feature selection using forward selection-partial least squares (FS-PLS) as previously described (11, 12). FS-PLS was performed separately for each of the comparisons used in DESeq2. For each comparison, the genes identified as SDE in the same comparison were used with the following further thresholds: BH $p$-values < 0.01, absolute log2 fold-change > 1, and base mean > 150.

The following FS-PLS parameters were used. The "$p$-value threshold" (maximum threshold for maximum likelihood estimation $p$-value for model performance) and "beam" (number of models ran in parallel) parameters were consistent for all comparisons at 0.001 and 20, respectively. The "max" parameter (the maximum number of features added) was chosen according to the number of groups contrasted against COVID-19: 2 for COVID-19 *vs*. bacterial and COVID-19 *vs*. viral, and 4 for COVID-19 *vs*. bacterial and viral combined, and COVID-19 *vs*. bacterial, viral, and non-infected but unwell controls combined. Different "max" parameters were chosen to reflect the different number of comparator groups in each model.

For each FS-PLS comparison, the top signature with the highest area under the receiver operating characteristic (ROC) curve (AUC) in the training set was selected and all genes were pooled to form a single signature. The performance of the signature was tested through combining the normalised gene counts of all genes in the signature into a single score (simple DRS), as in (12) and using the pROC package (13) to calculate the AUC for each signature identified. The coefficients calculated for each gene from generalised logistic regression models contrasting COVID-19 to the comparator group used for signature discovery by FS-PLS, which were multiplied by their respective gene's normalised counts to obtain a retrained disease risk score (DRS) for each sample as follows:

$$Retrained\ Disease\ Risk\ Score\ (DRS)^i = \sum gene\ expression_m^i \times gene\ weight_m$$

where *i* represents each sample and *m* represents each gene in the signature.

All statistical analyses were performed in R version 4.0.3 (Team 2020).(14)

**Validation cohorts used for RT-qPCR analysis**

**Figure S2**

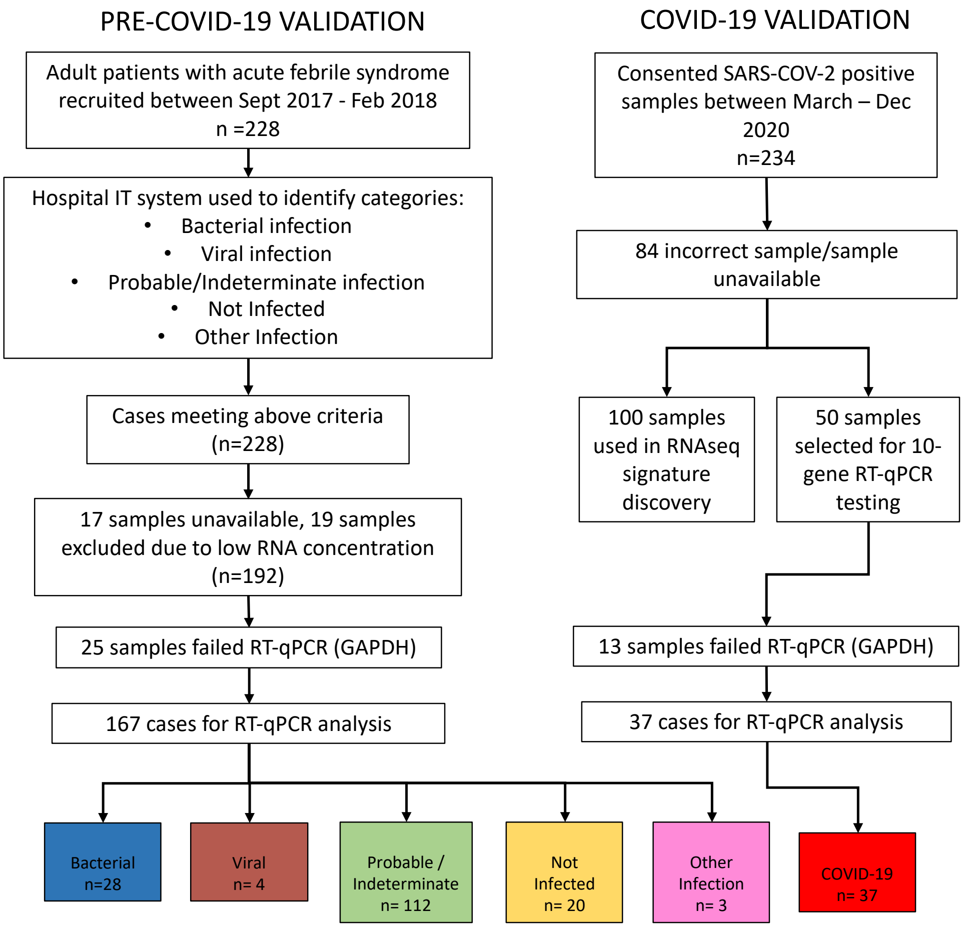


**Supplementary Figure S2**: **Study profile of pre-COVID-19 and COVID-19 validation cohorts**. Other infections were mycobacterial, fungal and parasitic. Pre-COVID-19 validation cohort were as described in (1).

**Prospective validation of the diagnostic gene signature**

Using RNA obtained from acute admission blood samples, the diagnostic gene signature was then validated using RT-qPCR. Exons within the genes included in the signature from the RNA-seq analysis were explored using exon counts to identify the optimal region of the gene to target in RT-qPCR validation. The exons' mean counts and AUCs for differentiating between COVID-19 *vs*. bacterial, viral, bacterial + viral, and bacterial + viral + not infected groups were evaluated and used to select the most promising transcripts to target for validation.

Genes identified in the signature discovery stage were taken forward to validation using RT-PCR. TaqMan assays were designed and developed in this study, and purchased from IDT Integrated DNA Technologies (IDT, Coralville, IA). Analytical sensitivity and specificity of the six assays were evaluated using human genomic DNA (Promega Corporation), respectively.

The signature performance was assessed using the Biomark HD (Fluidigm) and the 192.24 Dynamic Array™ integrated fluidic circuit (IFC) following manufacturer instructions. The experimental workflow consisted of 4 steps: reverse transcription, pre- amplification, on-chip gene expression, and data analysis.

Expression of genes in the diagnostic gene signature was quantified using RT-qPCR with *GAPDH* used as a housekeeping gene; the signature score was calculated using the simple DRS as described in the main methods of the manuscript, taking account of the inverse relation between Ct and transcript quantity. Samples from patients in the following phenotypic groups were included: COVID-19, Bacterial Infections, Viral Infections, Not Infected, Probable/Indeterminate, and Other Infection. Two replicates of each gene were used for each sample. Samples missing measurements for both replicates of *GAPDH* were removed. In order to have only one measurement per sample per gene for downstream analyses, the average across both replicates for each gene was taken for each sample. If one of the replicates was missing a measurement, the other replicate was used and if both replicates were missing, the value for downstream analyses was set to NA (except for *GAPDH*).

Optimal performance of the signature was calculated through retraining model weights using generalised logistic regression models to calculate the retrained DRS as described above. The retrained DRS and simple DRS were used to calculate the ROC curve and AUC.

***In silico* validation of the diagnostic gene signature**

A publicly available RNA-seq dataset (15) was used for *in silico* validation of the diagnostic gene signature. Rather than downloading the counts for the publicly available RNA-seq dataset, the raw fastq files were downloaded and the same pipeline was followed as described for the discovery datasets (batch A and B) to enable fair comparison. Fastq files from (15) were downloaded from Gene Expression Omnibus accession GSE161731 and were trimmed using trimmomatic (2). Quality control was performed using FastQC (3) and MultiQC (4), mapping to the human genome was carried out using STAR (5), and featureCounts (6) was used for read counting. Release 37 GENCODE GCh38 genome and annotation (7) were used. Raw counts were normalised using DESeq2 (8). The *in silico* RNA-seq validation set contained transcriptomic profiles from individuals infected with COVID-19, bacterial infections, influenza, seasonal coronaviruses and healthy controls. The performance of the host gene signature was explored using both the simple DRS and the retrained DRS using equations described above. The *in silico* RNA-seq dataset contained "early", "middle" and "late" samples from patients, representing time between sample collection and symptom onset (early ≤10 days, middle 11–21 days, late >21 days). As all blood RNA samples in our current study were obtained at the point of admission to hospital, we did not include the "late" group in our analysis. Calculations considered early COVID-19 cases alone as timings were similar to the BioAID cohort, as well as early and middle COVID-19 cases combined. Healthy controls were used for initial data pre-processing but not for signature performance evaluation. AUCs were calculated for: COVID-19 versus bacterial infections, COVID-19 versus viral infections (influenza and seasonal coronaviruses), and COVID-19 versus bacterial and viral infections combined.

### Supplementary Results

#### Signature discovery: Principal component analysis

18,247 quantified genes in the normalised, merged dataset were visualised on principal components 1 and 2 (Supplementary Fig. S3). COVID-19 samples overlapped with the other viral infection samples and, to a lesser degree, the bacterial and non-infected samples.

**Supplementary Figure S3: Principal component analysis of merged BioAID A and B RNAseq datasets**

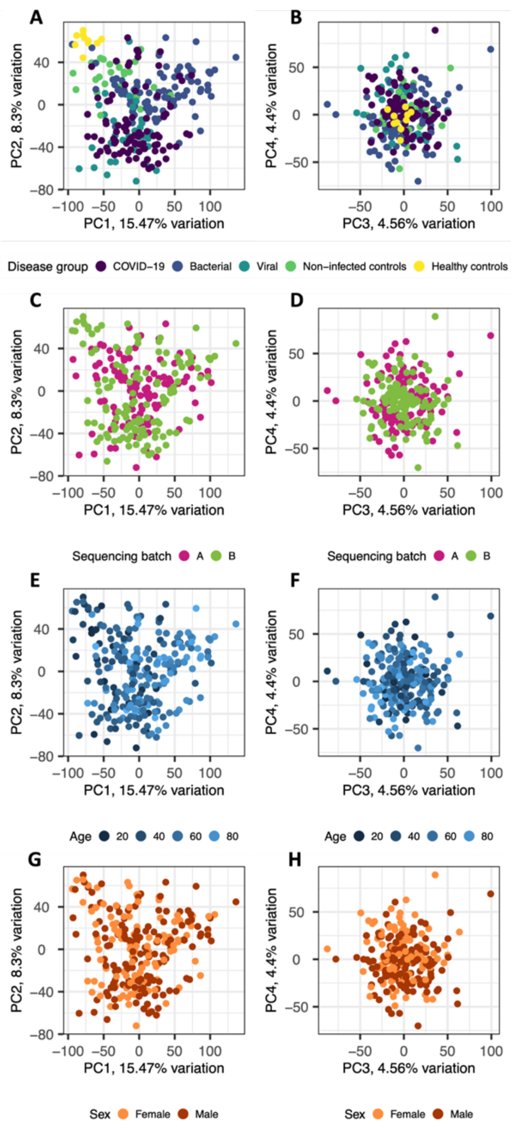


**Supplementary Figure S3:** Principal component analysis (PCA) plots with points representing samples and coloured according to disease group (A-B), sequencing batch (C-D), age in years (E-F), and sex (G-H) across either principal components (PC) 1-2 (A, C, E, G) or 3-4 (B, D, F, H).

Prior to identification of the optimal signature for differentiating between COVID-19 and the other disease groups, differential expression analysis was performed to reduce the search space. 7,726 genes were SDE (BH $p$-value < 0.05) between COVID-19 and bacterial infections, with 3,157 and 4,569 genes over- and under-expressed in COVID-19, respectively. 1,990 genes were SDE between COVID-19 and viral infections, with 776 and 1214 genes over- and under-expressed in COVID-19, respectively. There were 3,583 genes SDE between COVID-19 and non-infected controls, with 2,051 and 1,532 genes over- and under-expressed in COVID-19, respectively. When COVID-19 was contrasted to bacterial and viral infections combined, there were 4,889 SDE genes, with 1,825 and 3,064 genes over- and under-expressed in COVID-19, respectively. Finally, when COVID-19 was contrasted to all groups combined (bacterial, viral, and not infected), there were 3,811 SDE genes with 1,755 and 2,056 genes over- and under-expressed in COVID-19, respectively.

**Signature Discovery.**

**Supplementary Table S1 Genes included in two-gene to four-gene minimal signatures identified by FS-PLS for the 4 comparisons of interest.**

| Comparison | Signature (direction in COVID-19) | AUC (95% Confidence Interval) |
|---|---|---|
| COVID-19 *vs*. bacterial + viral + non-infected | *OASL* (up), *UPB1* (up), *IL1RN* (down), *ZNF684* (down) | 80.9% (75.4%-86.5%) |
| COVID-19 *vs*. bacterial + viral | *UPB1* (up), *ENTPD7* (down), *NFKBIE* (down), *CDKN1C* (down) | 86.2% (81.2%-91.1%) |
| COVID-19 *vs*. bacterial | *CD44* (down), *OTOF* (up) | 91.2% (87.0%-95.4%) |
| COVID-19 *vs*. viral | *MSR1* (down), *UPB1* (up) | 86.1% (78.5%-93.8%) |

AUC = area under the receiver operating characteristic (ROC) curve (AUC); CI = confidence interval. Gene abbreviations. *OASL*, 2'-5'-oligoadenylate synthetase-like protein; *UPB1*, Beta-ureidopropionase; *IL1RN*, interleukin-1 receptor antagonist protein; *ZNF684,* Zinc finger protein 684; *ENTPD7,* Ectonucleoside triphosphate diphosphohydrolase 7; *NFKBIE,* Nuclear factor of kappa light polypeptide gene enhancer in B-cells inhibitor, epsilon; *CDKN1C,* Cyclin-dependent kinase inhibitor 1C; *CD44,* CD44 cell surface antigen; *OTOF,* Otoferlin; *MSR1,* Macrophage scavenger receptor 1 (CD204).

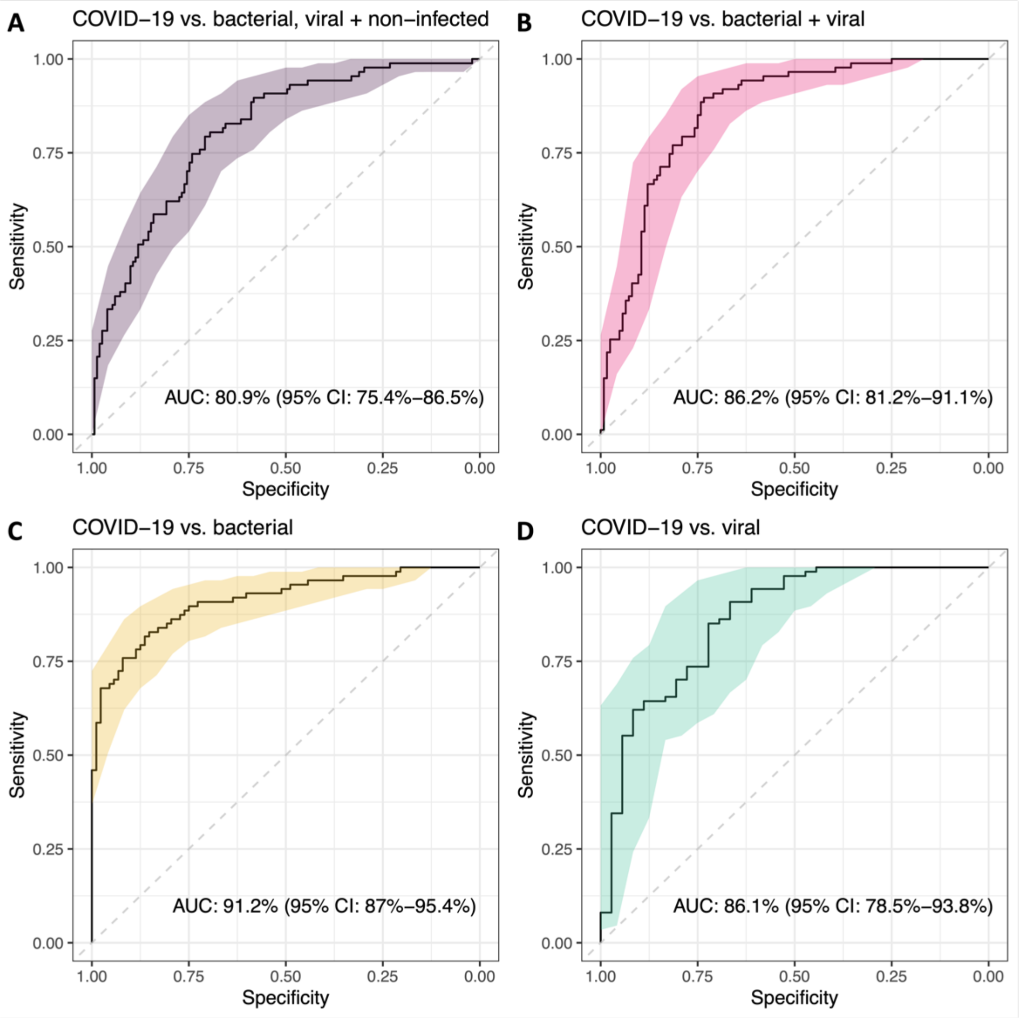


**Supplementary Figure S4. Performance of each of the two-gene or four-gene diagnostic gene signatures identified by FS-PLS in the discovery RNA-seq dataset** (see Supplementary Table S1): A) COVID-19 *vs*. bacterial, viral and not infected; B) COVID-19 *vs*. bacterial and viral infections; C) COVID-19 *vs*. bacterial infections; and D) COVID-19 *vs*. viral infections. AUCs shown by solid lines and 95% confidence intervals (CI) indicated by shaded areas.

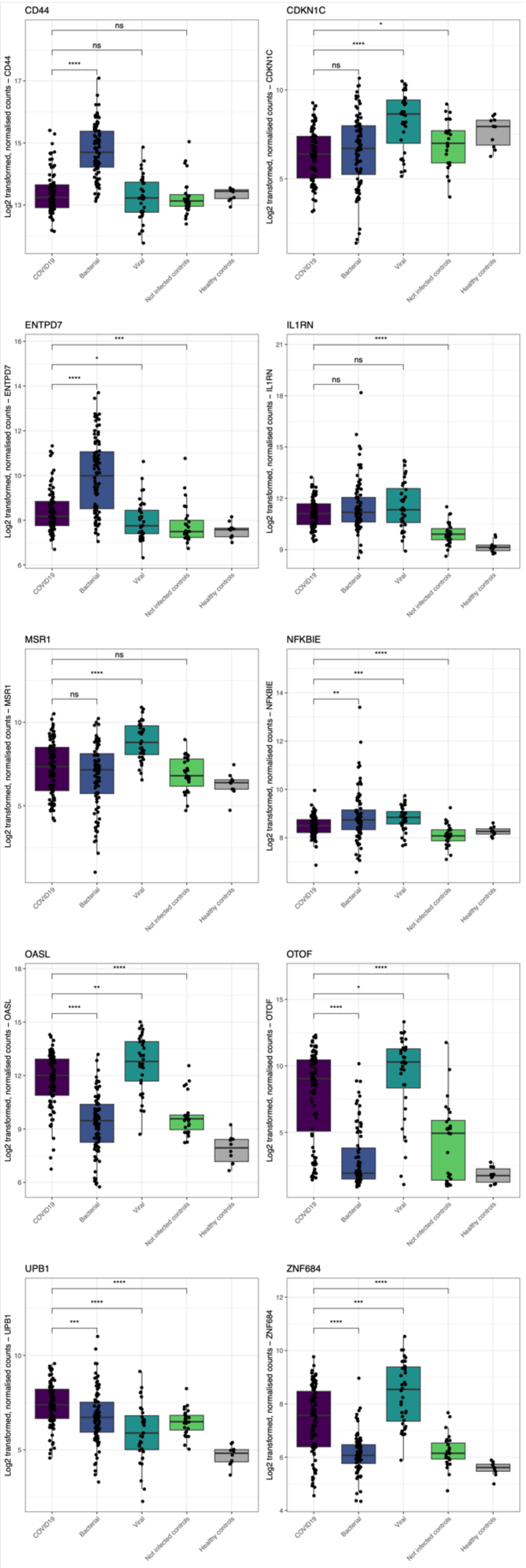


**Supplementary Figure S5: Boxplots for each of the 10 genes in the signature in the Discovery cohorts measured using RNA-seq**. Comparison of mean expression levels between COVID-19 and other disease groups are shown on each plot (****: $p \leq 0.0001$; ***: $p \leq 0.001$; **: $p \leq 0.01$; *: $p \leq 0.05$; ns, not significant).

### RT-qPCR Validation of 10-gene signature

### Supplementary Figure S6.

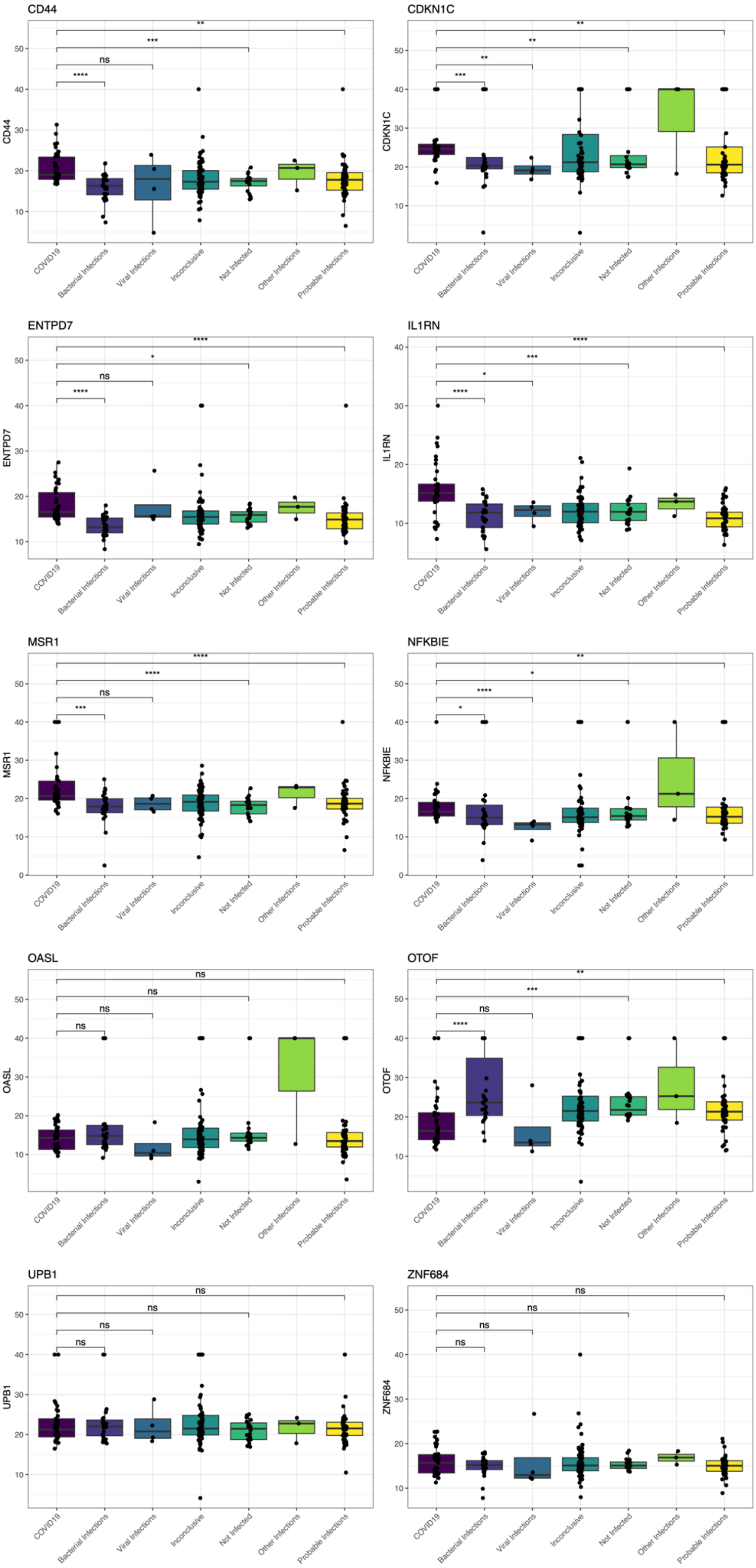

**Supplementary Figure S6:** Boxplots for each of the 10 genes in the signature measured using RT-qPCR in each validation group. Comparison of mean expression levels between COVID-19 and other disease groups are shown on each plot (****: $p \leq 0.0001$; ***: $p \leq 0.001$; **: $p \leq 0.01$; *: $p \leq 0.05$; ns, not significant).

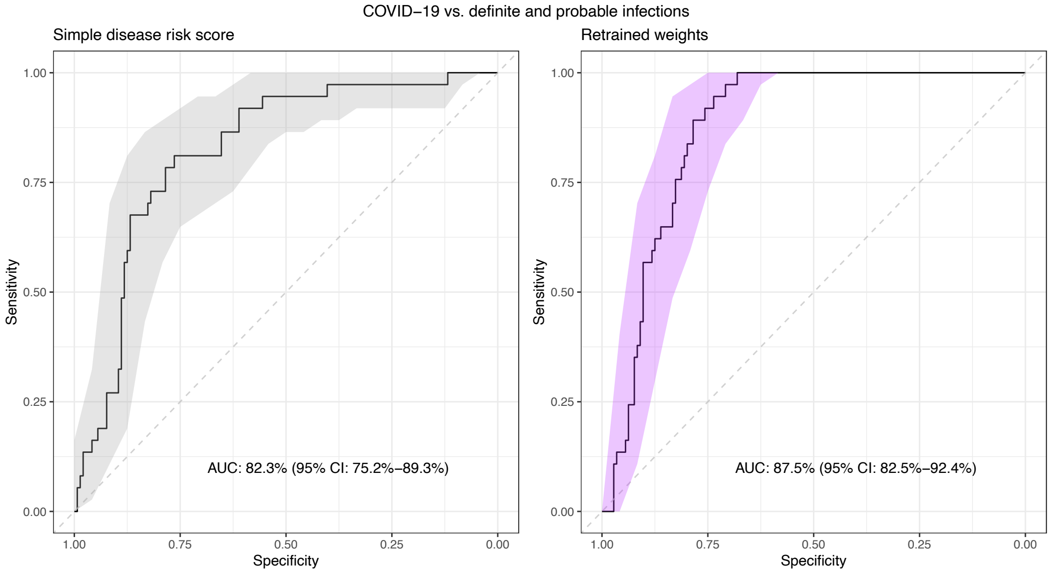


**Supplementary Figure S7**: **ROC curves showing the performance of the 10-gene signature in the RT-qPCR dataset in differentiating between COVID-19 and all other infection groups**. The performance of the 10-gene signature using the simple disease risk score is shown on the left-hand side panel, whilst the performance using retrained weights is shown in the right-hand side panel.

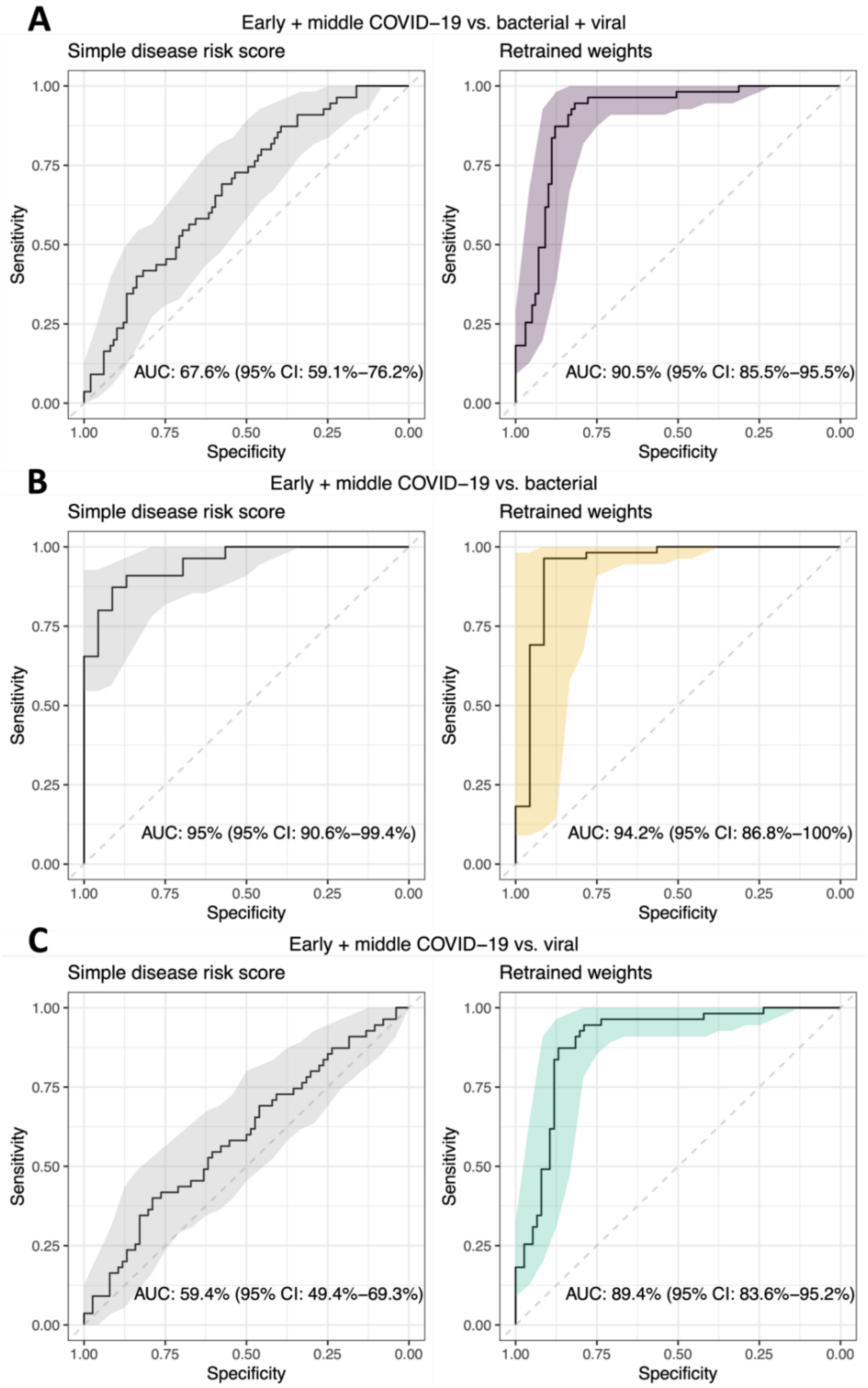


**Supplementary Figure S8: Performance of 10-gene signature in differentiating early and mid-duration COVID-19 from other infections.** ROC curves showing the performance of the 10-gene signature in the *in-silico* RNA-seq validation dataset in differentiating early and mid-duration COVID-19 from bacterial and viral infections combined (A), COVID-19 from bacterial infections (B), and COVID-19 from other viral infections (C). The performance of the 10-gene signature using the simple disease risk score is shown on the left-hand side panel, whilst the performance using retrained weights is shown in the right-hand side panel.

**Supplementary Table S2. Differentially expressed genes comparing Infected vs. Not-infected patients. Negative and positive log2 fold-change values indicate that the gene decreases or increases in infected patients *vs*. not infected patients.**

| | **BaseMean** | **log2FoldChange** | **padj** | **Gene** |
|---|---|---|---|---|
| ENSG00000078098 | 85.25386666 | -3.467043437 | 6.60E-20 | FAP |
| ENSG00000165185 | 880.1088042 | -1.906703107 | 6.60E-20 | KIAA1958 |
| ENSG00000151364 | 28.85883374 | -3.369172224 | 1.21E-18 | KCTD14 |
| ENSG00000155886 | 4.441784293 | 3.395759371 | 1.37E-18 | SLC24A2 |
| ENSG00000169245 | 323.8383143 | -3.86237307 | 2.80E-18 | CXCL10 |
| ENSG00000133101 | 111.487718 | -3.641010023 | 3.60E-17 | CCNA1 |
| ENSG00000226004 | 39.61546682 | -3.040011817 | 4.86E-16 | LINC02528 |
| ENSG00000225492 | 483.6946249 | -3.540359136 | 6.43E-16 | GBP1P1 |
| ENSG00000115267 | 5905.575906 | -2.122755474 | 1.15E-15 | IFIH1 |
| ENSG00000196123 | 154.4151851 | -1.95260772 | 1.15E-15 | KIAA0895L |
| ENSG00000121797 | 344.9332042 | -1.834702607 | 5.85E-15 | CCRL2 |
| ENSG00000163666 | 48.88648358 | -3.124325138 | 5.85E-15 | HESX1 |
| ENSG00000171658 | 44.44962135 | -2.219820344 | 8.11E-15 | NMRAL2P |
| ENSG00000254602 | 32.63364605 | -2.685633172 | 1.17E-14 | AP000662.1 |
| ENSG00000196141 | 1029.402028 | -2.467627446 | 1.40E-14 | SPATS2L |
| ENSG00000188313 | 8889.600279 | -1.529625145 | 1.84E-14 | PLSCR1 |
| ENSG00000002549 | 4178.241191 | -2.053178714 | 2.33E-14 | LAP3 |
| ENSG00000170439 | 153.8578903 | -2.797275512 | 2.33E-14 | METTL7B |
| ENSG00000279320 | 479.0848013 | -1.556935071 | 2.55E-14 | AC069528.2 |
| ENSG00000074410 | 31.43339765 | -2.398440306 | 3.02E-13 | CA12 |
| ENSG00000134809 | 534.3608265 | -1.939949247 | 3.60E-13 | TIMM10 |
| ENSG00000235321 | 74.31228657 | -1.851805803 | 4.11E-13 | AC007556.1 |
| ENSG00000283064 | 116.7255288 | -1.624403956 | 4.38E-13 | AL353759.1 |
| ENSG00000177409 | 17206.20415 | -1.706931709 | 4.48E-13 | SAMD9L |
| ENSG00000234232 | 29.20866474 | -1.985568519 | 5.03E-13 | AC243772.3 |
| ENSG00000135114 | 3896.757758 | -2.188768722 | 6.50E-13 | OASL |
| ENSG00000254420 | 131.1728795 | -2.747277697 | 1.05E-12 | AP003086.1 |
| ENSG00000134326 | 4787.626299 | -2.172647636 | 1.15E-12 | CMPK2 |
| ENSG00000110079 | 538.6664072 | -1.866188273 | 1.15E-12 | MS4A4A |
| ENSG00000118160 | 9.772597163 | -3.468290057 | 1.15E-12 | SLC8A2 |
| ENSG00000137628 | 7937.687895 | -1.889432008 | 1.18E-12 | DDX60 |
| ENSG00000116663 | 722.7288802 | -1.576934693 | 1.28E-12 | FBXO6 |
| ENSG00000224789 | 122.89286 | -2.312965666 | 1.28E-12 | AC012363.1 |
| ENSG00000163746 | 79.94162913 | -1.880712297 | 1.28E-12 | PLSCR2 |
| ENSG00000078081 | 291.2807663 | -2.802215584 | 1.28E-12 | LAMP3 |
| ENSG00000089127 | 8023.083035 | -2.118505342 | 1.35E-12 | OAS1 |
| ENSG00000068079 | 1774.800706 | -1.67353107 | 1.55E-12 | IFI35 |

| | | | | |
|---|---|---|---|---|
| ENSG00000140464 | 2547.551813 | -1.726610978 | 1.78E-12 | PML |
| ENSG00000184979 | 644.095283 | -2.948405892 | 1.83E-12 | USP18 |
| ENSG00000136514 | 1032.534983 | -2.152307133 | 2.13E-12 | RTP4 |
| ENSG00000225964 | 277.2645817 | -1.917979953 | 2.27E-12 | NRIR |
| ENSG00000155363 | 1498.905589 | -1.690995811 | 2.47E-12 | MOV10 |
| ENSG00000107201 | 9533.915957 | -1.682038596 | 2.47E-12 | DDX58 |
| ENSG00000117010 | 186.2337314 | -1.595310329 | 2.79E-12 | ZNF684 |
| ENSG00000111335 | 12799.78896 | -2.126271558 | 2.80E-12 | OAS2 |
| ENSG00000238015 | 74.42075567 | -2.522589875 | 4.12E-12 | AC104837.2 |
| ENSG00000172159 | 891.7769514 | -1.530149503 | 4.93E-12 | FRMD3 |
| ENSG00000183831 | 20.46950819 | -2.799523068 | 5.71E-12 | ANKRD45 |
| ENSG00000117226 | 2091.485578 | -1.69531389 | 7.02E-12 | GBP3 |
| ENSG00000255221 | 761.0115997 | -2.259641093 | 7.56E-12 | CARD17 |
| ENSG00000133106 | 5548.85876 | -1.982732933 | 7.56E-12 | EPSTI1 |
| ENSG00000117228 | 12280.34862 | -2.095237655 | 8.09E-12 | GBP1 |
| ENSG00000152766 | 1142.445428 | -2.109783836 | 8.29E-12 | ANKRD22 |
| ENSG00000108387 | 268.0976844 | -2.17869545 | 8.29E-12 | SEPTIN4 |
| ENSG00000217159 | 26.35620514 | -1.691818653 | 8.82E-12 | LARP1P1 |
| ENSG00000255397 | 17.8626076 | -2.024862333 | 1.03E-11 | NASPP1 |
| ENSG00000265566 | 14.69862431 | -2.079731788 | 1.07E-11 | RN7SL605P |
| ENSG00000059378 | 3297.977833 | -1.640778615 | 1.08E-11 | PARP12 |
| ENSG00000162772 | 157.1889385 | -2.088337891 | 1.37E-11 | ATF3 |
| ENSG00000233646 | 21.05874329 | -1.815010195 | 1.56E-11 | OR52T1P |
| ENSG00000254649 | 19.31058126 | -2.913647465 | 1.58E-11 | AP003086.2 |
| ENSG00000143891 | 880.8593516 | -1.596013352 | 2.90E-11 | GALM |
| ENSG00000173193 | 30984.35632 | -1.533341148 | 3.07E-11 | PARP14 |
| ENSG00000108771 | 1115.156826 | -1.904705805 | 3.49E-11 | DHX58 |
| ENSG00000124256 | 2997.576971 | -1.577325408 | 4.09E-11 | ZBP1 |
| ENSG00000152778 | 3944.479448 | -1.552614837 | 4.16E-11 | IFIT5 |
| ENSG00000163568 | 1685.394267 | -1.558660831 | 4.61E-11 | AIM2 |
| ENSG00000134321 | 20037.98293 | -2.552480472 | 4.79E-11 | RSAD2 |
| ENSG00000205837 | 24.9218279 | -2.319902331 | 5.09E-11 | LINC00487 |
| ENSG00000119922 | 32272.07226 | -2.087338827 | 7.07E-11 | IFIT2 |
| ENSG00000158488 | 5.897594986 | 1.765603328 | 1.22E-10 | CD1E |
| ENSG00000233785 | 37.44734182 | -2.112776466 | 1.24E-10 | AC131011.1 |
| ENSG00000138646 | 6553.1179 | -2.250907618 | 1.31E-10 | HERC5 |
| ENSG00000138642 | 1730.733041 | -1.651073554 | 1.43E-10 | HERC6 |
| ENSG00000221955 | 23.14283836 | -1.501976343 | 1.44E-10 | SLC12A8 |
| ENSG00000223387 | 50.01018771 | -2.549081965 | 1.44E-10 | LINC02068 |
| ENSG00000132530 | 11345.16585 | -1.834541779 | 1.50E-10 | XAF1 |
| ENSG00000142408 | 18.55732015 | 2.157123776 | 1.57E-10 | CACNG8 |
| ENSG00000137965 | 6380.290887 | -2.160854979 | 1.81E-10 | IFI44 |
| ENSG00000269720 | 82.42821876 | -2.258568554 | 2.04E-10 | CCDC194 |

| | | | | |
|---|---|---|---|---|
| ENSG00000226751 | 27.24670746 | -1.907754486 | 2.51E-10 | NA |
| ENSG00000259641 | 7.893466298 | 1.612525908 | 2.53E-10 | NA |
| ENSG00000198785 | 92.01533994 | -2.139902776 | 2.99E-10 | GRIN3A |
| ENSG00000143344 | 321.9125707 | -1.637875863 | 3.21E-10 | RGL1 |
| ENSG00000175518 | 33.75470294 | -1.673935784 | 3.34E-10 | UBQLNL |
| ENSG00000280248 | 302.2209862 | -1.625532267 | 3.38E-10 | AC124319.3 |
| ENSG00000260096 | 5.605742866 | -2.200025837 | 5.56E-10 | DNM1P33 |
| ENSG00000277511 | 32.11440393 | -1.604997258 | 5.82E-10 | AC116407.2 |
| ENSG00000108950 | 330.0485822 | -1.86689792 | 5.88E-10 | FAM20A |
| ENSG00000157601 | 18017.80955 | -2.086221493 | 5.90E-10 | MX1 |
| ENSG00000102794 | 16.17617427 | -2.419208182 | 6.23E-10 | ACOD1 |
| ENSG00000179639 | 160.3502663 | 1.795039581 | 6.30E-10 | FCER1A |
| ENSG00000277301 | 11.20138225 | 1.631593446 | 8.07E-10 | AL034550.2 |
| ENSG00000198744 | 4.696190786 | 2.352874381 | 8.14E-10 | MTCO3P12 |
| ENSG00000119917 | 23855.32651 | -2.079746035 | 8.21E-10 | IFIT3 |
| ENSG00000108700 | 21.79282947 | -2.564504907 | 9.04E-10 | CCL8 |
| ENSG00000188282 | 138.2551851 | -1.869637644 | 9.45E-10 | RUFY4 |
| ENSG00000144730 | 5.317907675 | -2.155164253 | 9.58E-10 | IL17RD |
| ENSG00000224295 | 43.23823315 | -1.573312185 | 1.01E-09 | OLFM5P |
| ENSG00000251072 | 6.957779784 | -1.819441979 | 1.11E-09 | LMNB1-DT |
| ENSG00000120217 | 2412.213341 | -1.835382616 | 1.13E-09 | CD274 |
| ENSG00000175643 | 167.0089801 | -1.75766374 | 1.38E-09 | RMI2 |
| ENSG00000258159 | 5.877138168 | -2.086400385 | 1.42E-09 | IMMP1LP2 |
| ENSG00000228863 | 249.9722004 | -2.415427657 | 1.61E-09 | AL121985.1 |
| ENSG00000258227 | 772.1821377 | -1.543573868 | 1.65E-09 | CLEC5A |
| ENSG00000111331 | 18191.49514 | -2.183157752 | 1.72E-09 | OAS3 |
| ENSG00000199668 | 8.111239971 | -2.198287865 | 2.17E-09 | Y_RNA |
| ENSG00000139572 | 925.040975 | -2.153459913 | 2.18E-09 | GPR84 |
| ENSG00000237927 | 170.7399332 | -1.713573715 | 2.21E-09 | AL078604.2 |
| ENSG00000137869 | 59.55260167 | -2.436809017 | 2.21E-09 | CYP19A1 |
| ENSG00000248553 | 13.08809783 | -1.560767404 | 2.36E-09 | OR52H2P |
| ENSG00000274376 | 9.593941149 | 2.181238621 | 2.36E-09 | ADAMTS7P1 |
| ENSG00000121380 | 18.71007898 | -1.906622456 | 2.82E-09 | BCL2L14 |
| ENSG00000168062 | 428.879651 | -2.161535869 | 2.93E-09 | BATF2 |
| ENSG00000229534 | 22.08686628 | -1.59314146 | 3.33E-09 | HNRNPA1P53 |
| ENSG00000177294 | 101.7678519 | -2.049198326 | 3.53E-09 | FBXO39 |
| ENSG00000135903 | 9.312067561 | -2.447593866 | 4.09E-09 | PAX3 |
| ENSG00000213557 | 6.812995224 | 1.552224598 | 4.49E-09 | AC068050.1 |
| ENSG00000108691 | 45.5056566 | -2.703344672 | 4.56E-09 | CCL2 |
| ENSG00000163016 | 79.37628969 | -2.353825768 | 6.02E-09 | ALMS1P1 |
| ENSG00000123610 | 2914.852129 | -1.605858071 | 6.02E-09 | TNFAIP6 |
| ENSG00000183134 | 100.4547601 | 1.855553109 | 6.38E-09 | PTGDR2 |
| ENSG00000249173 | 143.1131672 | -2.204379641 | 6.70E-09 | LINC01093 |

| | | | | |
|---|---|---|---|---|
| ENSG00000246363 | 20.43973863 | 1.555472988 | 7.12E-09 | LINC02458 |
| ENSG00000224666 | 15.97051242 | -2.091081966 | 9.57E-09 | ETV7-AS1 |
| ENSG00000149131 | 1852.147142 | -2.104525113 | 1.06E-08 | SERPING1 |
| ENSG00000234493 | 10.43033436 | 1.957218822 | 1.07E-08 | RHOXF1P1 |
| ENSG00000283399 | 20.56589169 | 1.508630868 | 1.17E-08 | AC004381.3 |
| ENSG00000126709 | 5996.143229 | -1.92331431 | 1.24E-08 | IFI6 |
| ENSG00000250696 | 51.39903105 | 2.099561055 | 1.38E-08 | AC111000.4 |
| ENSG00000125355 | 50.76431018 | -2.008404901 | 1.99E-08 | TMEM255A |
| ENSG00000091181 | 192.167867 | 1.979299259 | 2.33E-08 | IL5RA |
| ENSG00000246375 | 12.94838253 | -2.03230096 | 2.66E-08 | PPM1K-DT |
| ENSG00000154099 | 22.61288793 | -2.906325014 | 2.75E-08 | DNAAF1 |
| ENSG00000265531 | 686.3567053 | -1.672652133 | 3.13E-08 | FCGR1CP |
| ENSG00000154764 | 18.29411941 | 1.540755515 | 3.44E-08 | WNT7A |
| ENSG00000183691 | 32.12337821 | 1.727259882 | 3.47E-08 | NOG |
| ENSG00000137959 | 14414.18079 | -2.199601806 | 3.58E-08 | IFI44L |
| ENSG00000204455 | 7.501416094 | 1.515089333 | 3.66E-08 | TRIM51BP |
| ENSG00000189325 | 8.834169178 | -2.04068392 | 3.73E-08 | BNIP5 |
| ENSG00000168026 | 224.8044766 | -1.595989309 | 4.61E-08 | TTC21A |
| ENSG00000185745 | 15065.94677 | -2.215862914 | 4.64E-08 | IFIT1 |
| ENSG00000206199 | 23.24959608 | 1.507856423 | 6.72E-08 | ANKUB1 |
| ENSG00000255355 | 21.02610312 | -2.018387942 | 6.82E-08 | AP000640.2 |
| ENSG00000234756 | 4.918674239 | -1.965115767 | 7.90E-08 | LINC02621 |
| ENSG00000258872 | 90.00594367 | -1.547178142 | 7.91E-08 | FDPSP3 |
| ENSG00000010030 | 679.7936231 | -2.043205643 | 8.93E-08 | ETV7 |
| ENSG00000162654 | 5388.33776 | -1.550943593 | 9.22E-08 | GBP4 |
| ENSG00000280270 | 22.64028696 | -1.904226828 | 9.56E-08 | AC068299.2 |
| ENSG00000178175 | 105.8979255 | -1.535371209 | 9.71E-08 | ZNF366 |
| ENSG00000163081 | 5.697358542 | -1.931823623 | 9.71E-08 | CCDC140 |
| ENSG00000280007 | 54.4823232 | -1.748808417 | 9.76E-08 | AC008079.1 |
| ENSG00000121900 | 8.913957284 | -1.970624501 | 9.76E-08 | TMEM54 |
| ENSG00000283648 | 3.944403569 | -1.675962244 | 1.14E-07 | AC006974.2 |
| ENSG00000260325 | 4.840053989 | -2.173990572 | 1.22E-07 | HSPB9 |
| ENSG00000230013 | 5.986222485 | -1.536585771 | 1.24E-07 | CT70 |
| ENSG00000138395 | 6.71210574 | 1.757279843 | 1.24E-07 | CDK15 |
| ENSG00000130558 | 13.91645622 | 1.785324982 | 1.25E-07 | OLFM1 |
| ENSG00000173114 | 78.70959495 | 1.632239422 | 1.34E-07 | LRRN3 |
| ENSG00000103355 | 29.60136375 | 2.374727328 | 1.34E-07 | PRSS33 |
| ENSG00000280276 | 25.90712667 | -2.197505695 | 1.38E-07 | AC009229.4 |
| ENSG00000226500 | 4.288575927 | -1.843521278 | 1.84E-07 | AC243772.1 |
| ENSG00000165949 | 4467.370915 | -2.459906715 | 1.88E-07 | IFI27 |
| ENSG00000233975 | 7.124862613 | -1.813204701 | 1.91E-07 | LINC02574 |
| ENSG00000211961 | 6.501356078 | -2.543078646 | 2.04E-07 | IGHV1-45 |
| ENSG00000227292 | 17.12454533 | -2.548325465 | 2.26E-07 | AC009229.1 |

| | | | | |
|---|---|---|---|---|
| ENSG00000232884 | 9.221569496 | -1.5570603 | 2.63E-07 | AF127936.1 |
| ENSG00000211972 | 16.64737185 | -1.793044132 | 2.98E-07 | IGHV3-66 |
| ENSG00000161133 | 6.9783616 | -2.181145635 | 3.35E-07 | USP41 |
| ENSG00000227238 | 3.998280881 | -1.719435281 | 4.17E-07 | TTC39DP |
| ENSG00000165507 | 39.14821104 | -1.737316148 | 4.23E-07 | DEPP1 |
| ENSG00000088836 | 5.245161653 | -1.621262141 | 4.42E-07 | SLC4A11 |
| ENSG00000227502 | 30.25618843 | -1.751862272 | 5.04E-07 | MROCKI |
| ENSG00000180251 | 17.8132496 | -1.71573513 | 5.94E-07 | SLC9A4 |
| ENSG00000197646 | 118.3498231 | -1.633151018 | 7.41E-07 | PDCD1LG2 |
| ENSG00000282850 | 3.348959768 | 1.556673046 | 7.49E-07 | RHOXF1P2 |
| ENSG00000236712 | 5.260132486 | -1.948087521 | 8.39E-07 | AC079448.1 |
| ENSG00000164825 | 3.939067768 | -1.845014015 | 9.10E-07 | DEFB1 |
| ENSG00000079215 | 672.2343468 | -1.731318611 | 1.03E-06 | SLC1A3 |
| ENSG00000105366 | 31.57798081 | 2.196529471 | 1.10E-06 | SIGLEC8 |
| ENSG00000168658 | 9.318173255 | -1.571964178 | 1.22E-06 | VWA3B |
| ENSG00000054179 | 6.083500249 | 1.776975191 | 1.22E-06 | ENTPD2 |
| ENSG00000227017 | 7.415382506 | -1.64624464 | 1.30E-06 | AC007036.1 |
| ENSG00000105205 | 744.9141093 | 1.528681629 | 1.30E-06 | CLC |
| ENSG00000183347 | 209.9398803 | -1.719734526 | 1.63E-06 | GBP6 |
| ENSG00000249310 | 2.924705584 | -1.629934605 | 1.66E-06 | APOBEC3B-AS1 |
| ENSG00000134765 | 29.79566289 | 1.535206116 | 1.72E-06 | DSC1 |
| ENSG00000147647 | 4.93127958 | -1.560416993 | 1.81E-06 | DPYS |
| ENSG00000088827 | 3848.737654 | -2.083899219 | 1.91E-06 | SIGLEC1 |
| ENSG00000239887 | 26.81480018 | -1.506365927 | 2.03E-06 | C1orf226 |
| ENSG00000126562 | 3.902685864 | -1.657723981 | 2.57E-06 | WNK4 |
| ENSG00000154451 | 22170.36849 | -1.520769047 | 2.63E-06 | GBP5 |
| ENSG00000084734 | 8.597987484 | -1.657580066 | 4.53E-06 | GCKR |
| ENSG00000164070 | 49.69589403 | -1.701676036 | 5.75E-06 | HSPA4L |
| ENSG00000167601 | 44.97644515 | -1.828172241 | 6.19E-06 | AXL |
| ENSG00000211962 | 114.028337 | -1.722338973 | 6.48E-06 | IGHV1-46 |
| ENSG00000229160 | 15.79410091 | -1.850320966 | 7.51E-06 | AC009229.2 |
| ENSG00000156113 | 281.6471054 | -2.092033347 | 7.76E-06 | KCNMA1 |
| ENSG00000205927 | 17.4826888 | 1.744830258 | 7.76E-06 | OLIG2 |
| ENSG00000154736 | 32.86050409 | 1.682936972 | 7.83E-06 | ADAMTS5 |
| ENSG00000105707 | 49.00045202 | -1.556006247 | 7.92E-06 | HPN |
| ENSG00000243144 | 18.07037609 | -1.958992266 | 1.36E-05 | AC079760.2 |
| ENSG00000131203 | 266.6409923 | -1.707689123 | 2.06E-05 | IDO1 |
| ENSG00000153246 | 3.780878958 | -1.856182728 | 2.13E-05 | PLA2R1 |
| ENSG00000211659 | 245.0645023 | -1.579355177 | 2.14E-05 | IGLV3-25 |
| ENSG00000161905 | 83.02248845 | 1.95037577 | 2.33E-05 | ALOX15 |
| ENSG00000118113 | 4430.313652 | -1.752415855 | 4.14E-05 | MMP8 |
| ENSG00000183578 | 37.76296381 | -1.532977994 | 4.58E-05 | TNFAIP8L3 |
